# De Novo Design of Ultrahigh-Affinity Miniproteins Targeting PD-L1 for Non-Invasive Imaging: Preclinical Validation and First-in-Human Study

**DOI:** 10.1101/2025.08.08.25333138

**Authors:** Lei Zhao, Fan Zhang, Renqi Yao, Siqi Hao, Gehan Zhang, Jinping Tao, Ge Guo, Xin Li, Guangming Yang, Tao Li, Hanlu Li, Saijia Cui, Fei Wang, Qi Wang, Zidan Song, Jiannan Feng, Beifen Shen, Yongming Yao, Yu Guo, Hua Zhu, Yi Hu

## Abstract

De novo protein design is becoming a powerful tool for diagnostic and therapeutic agent development. However, direct evidence remains lacking for dynamic binding of de novo designed binders both *in vivo* and clinical settings. PD-L1, quantified via immunohistochemistry, represents the most widely validated, utilized, and accepted biomarker for PD-1/PD-L1 therapy. Due to the spatial heterogeneity and temporal dynamics of PD-L1 expression, this molecule represents an ideal target for exploring the dynamic binding of de novo-designed binders. Herein, we report de novo-designed miniprotein PD-L1-3—an ultrahigh-affinity binder targeting the PD-1/PD-L1 interface with sub-picomolar affinity and hyper-stability. Preclinical and first-in-human studies demonstrated that this de novo-designed ultrahigh-affinity binder exhibits significantly improved binding specificity, enhanced tissue penetration, prolonged target-positive tumor retention and a favorable safety profile *in vivo* and in patients, enabling advanced diagnostic and therapeutic agent development.

## Main

De novo protein design has revolutionized biomolecular engineering^1–3^, enabling the creation of precision binders for previously "undruggable" targets through computational frameworks such as Rosetta^4^, RFdiffusion^5^ and ProteinMPNN^6^. These advances have yielded remarkable *in vitro* successes—generating ultra-stable mini-proteins against influenza hemagglutinin^7^ and computationally engineered cytokines with tunable immune activity^8^. Yet beneath this technical triumph lies a fundamental, unresolved challenge: the paucity of direct evidence for dynamic binding behavior of de novo designed proteins in both physiological environments and clinical contexts. This gap critically undermines translational potential, as *in vitro* binding kinetics rarely recapitulate the reality of living systems. The core contradiction emerges in two dimensions. First, current validation relies heavily on static structural techniques (cryo-EM^9^, X-ray crystallography^10^) and purified-system assays (SPR^11^, BLI^12^), which capture only equilibrium states—not the transient, fluctuating interactions governing *in vivo* efficacy. A binder exhibiting picomolar affinity against isolated targets may fail catastrophically when confronted with serum protein interference, pH gradients in tumor microenvironments^13^, or shear stress in vascular compartments^14^. Second, de novo designs prioritize thermodynamic stability over functional dynamics, often producing rigid scaffolds incapable of the conformational flexibility required for adaptive binding within densely packed cellular milieus^15^. Utilizing PET-CT imaging enables dynamic quantification of tissue distribution and tracking of real-time binding dynamics of de novo-designed proteins in living subjects^16^.

Growing evidence has shown the geographical heterogeneity^17^ and the dynamic changes in PD-L1 expression over time as the tumor and its immune microenvironment evolve^18^. More critically, interferons^19^, toll-like receptor ligands^20^, radiotherapy^21^, chemotherapy^22,23^, and targeted therapy^24^ have all been shown to affect the expression of PD-L1. All these data indicated that PD-L1 could be regarded as an ideal target for evaluating the dynamic binding of de novo-designed binder *in vivo* and in clinical settings.

De novo protein design approaches enable the targeted design of binders with high binding affinity to sites of interest^5,25–27^. Without any experimental optimization, the physically based Rosetta approach could directly design de novo binders with binding affinities ranging from 100 nM to 2 µM^28^. After further optimization through site saturation mutagenesis library (SSM) and combinatorial library screenings, binders with exceptionally high binding affinities ranging from 100 pM to 10 nM can be obtained^25,28^. Recently, there has been considerable progress in the de novo binder design using deep learning methods^5,6,26,29–31^. The deep network hallucination^26,31^ or RFdiffusion^5^, combined with ProteinMPNN^6^ could directly design de novo binders with higher binding affinity, ranging from 27 nM to 1.4 µM. In this work, we describe a design strategy that enables direct de novo design of an ultrahigh-affinity (1.15 pM) binder targeting PD-L1 eliminating the need for affinity optimization. Preclinical and first-in-human studies were performed to further reveal superior binding specificity, deeper tissue penetration, and optimized safety profiles of the ultrahigh-affinity de novo binder, demonstrating the promising clinical applicability of the de novo designed binder.

### De novo design of ultrahigh-affinity miniprotein targeting PD-L1 without subsequent experimental affinity optimization

We set out to directly design de novo protein binders with ultra-high binding affinity against the interface of PD-L1/PD-1. The binders were designed entirely from scratch without relying on the known PD-L1/PD-1 interactions (Fig. 1a). PatchDock^32^ achieved direct low-resolution shape matching with in silico miniprotein libraries^33,34^, and it was followed by a grid-based refinement of the rigid body orientation in the Rotamer interaction field (RIF)^35^. We first generated the RIF by docking billions of individual disembodied amino acids to the PD-L1/PD-1 interface we selected. Side-chain rotamer conformations are grown backwards for the placement of side-chain R-groups, and the backbone coordinates and target binding energies of the billions of typical amino acids that facilitate favourable hydrogen or nonpolar interactions are stored in a six-dimensional spatial hash table for quick look-up. Then, using a branch-and-bound searching algorithm, the protein scaffold library was docked into the field of the inverse rotamers, starting from low-resolution spatial grids to high-resolution spatial grids. This approach allows for quick approximation of the target interaction energy achievable by protein scaffolds docked against the region of the target we are interested in, based solely on scaffold backbone coordinates^25,28^.

**Figure 1.**
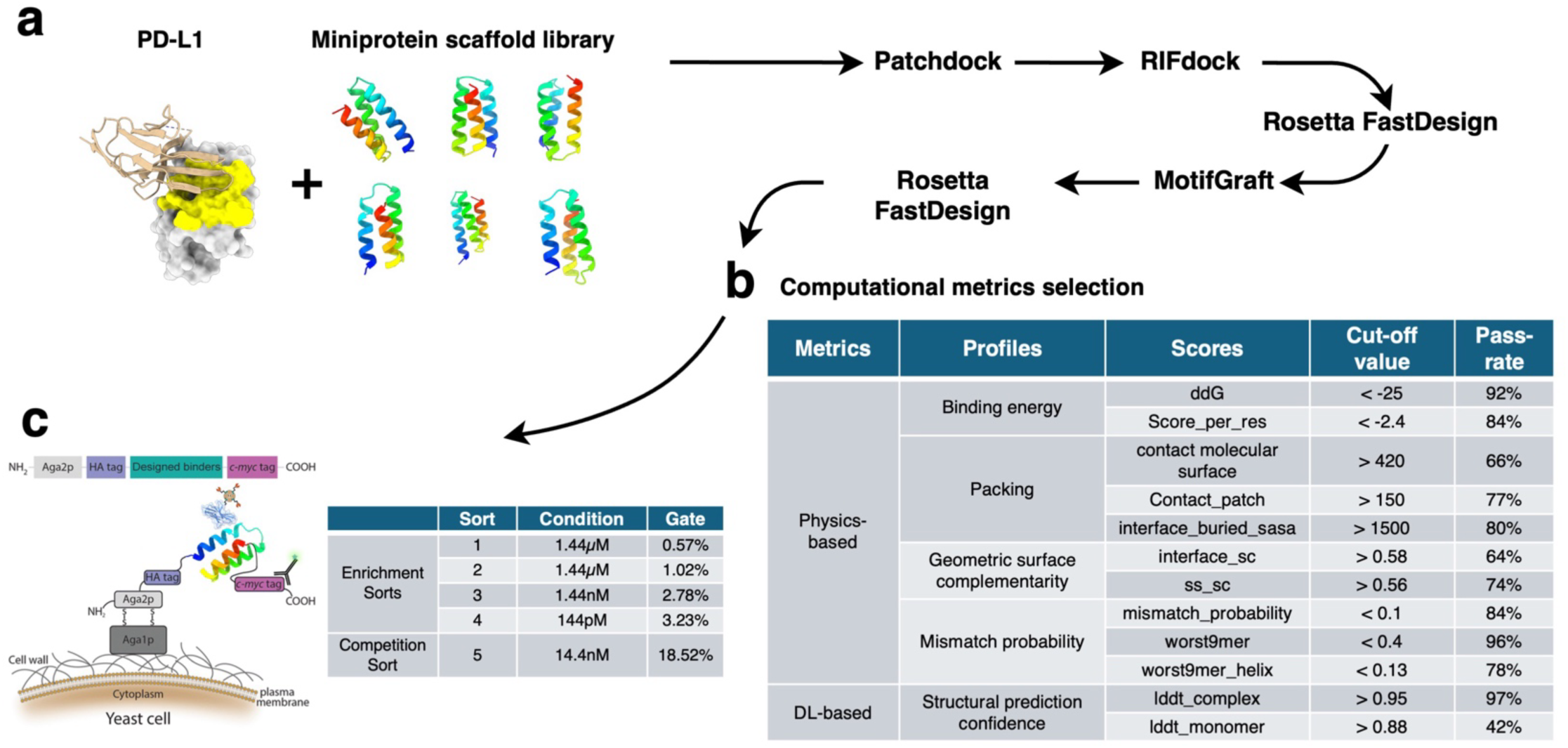
Overview of the computational design approach. **a,** The binding interface (yellow) of PD-1/PD-L1 was selected for de novo binder design. Patchdock and subsequent RIFdock were performed to identify shape and chemically complementary binding modes. The designs were further optimized utilizing an alternative Rosetta FastDesign protocol. To further refine the designed binders and broaden the exploration of possibilities, the motifs were extracted from the cream of the designed interfaces and subsequently integrated onto the scaffold library employing MotifGraft mover. The Motif-Grafted binders were further polished by the alternative Rosetta FastDesign protocol. **b,** Designs are then selected for experimental characterization based on physics-based and DL-based computational metrics. **c,** The top designs were synthesized and expressed on yeast cell surface. After 4 enrichment sorts and competition sort, the de novo designed binders with high binding affinities were identified.

The binders were further optimized using the Rosetta FastDesign protocol with several improvements, as previously described^25,28^. The binder motifs that had good interactions with the target PD-L1 were extracted and clustered. The best motif from each cluster was selected and then used to guide the protein scaffold library to superimpose onto it using MotifGraft mover. Subsequently, the binders were final optimized using the Rosetta FastDesign protocol above.

The advanced deep learning (DL) methods AlphaFold (AF)^36,37^ and RoseTTFold (RF)^38^ have achieved unparalleled accuracy in protein structure prediction. Up to now, AF and RF have been capable of accurately predicting the structures of naturally occurring proteins^39^. However, AF and RF generally require multiple sequence alignments, which contain rich co-evolutionary information about the residues of these naturally occurring proteins. Compared with proteins evolved from natural evolution, the sequence similarity of the de novo designed proteins that were previously reported^25,40,41^ is less than 30%. DeepAccuracyNet-Bert (DAN-Bert), which has achieved state-of-the-art performance in accuracy prediction in CASP14^42^, is capable of estimating per-residue accuracy and residue-residue distance signed error in protein models, even without any evolutionary alignments. Furthermore, DAN-Bert excels in speed, taking only about 0.5 GPU seconds per monomer^43^. This is approximately 10 times faster than the time AF takes for structure predictions. Therefore, the predicted global Cβ local distance difference test (l-DDT) of all the designed binders and their complexes were assessed by DAN-Bert, which are respectively referred to as lddt_monomer and lddt_complex.

The de novo-designed binders were evaluated using physics-based and DL-based metrics (Fig. 1b). In physics-based metrics, Rosetta was utilized for calculating the Rosetta binding energy (ddG) and scoring per residue (score_per_res), analyzing packing profiles that include contact molecular surface, contact molecular surface to critical hydrophobic residues (contact_patch), and the interface-buried contact molecular surface (interface_buried_sasa) during binder-target complex formation. It also includes geometric surface complementarity profiles covering the interface (interface_sc) and protein secondary structure elements (ss_sc). Mismatch probability profiles feature the geometric average of the probability of acquiring an incorrect secondary structure type at each position (mismatch_probability), and the structural alignment of 9-amino acid stretches from the helices (worst9mer_helix) or the entire sequence (worst9mer) of designed binders against fragments from the PDB^44,45^. In DL-based metrics, the structural prediction confidence of designed binders (lddt_monomer) and their protein complexes (lddt_complex) was assessed using DAN-Bert (Fig. 1b)^42^. The designs with the most favourable physics-based and DL-based metrics were selected for experimental validation (Fig. 1b and Extended Data Fig. 1).

### In vitro characterization

10^5^ designed binders were encoded into oligonucleotides and cloned into a yeast surface-expression vector for binder display (Fig. 1c). Using a fluorescently labeled PD-L1, these were enriched by 4 sequential rounds of fluorescence-activated cell sorting (FACS) (Fig. 1c and Extended Data Fig. 2). As shown in Extended Data Fig. 2d, even at 14.4 pM of PD-L1 target protein, detectable binding persisted. Yeast displaying high-affinity binders were harvested after incubation at 144 pM (Extended Data Fig. 2d). To verify tight binding at PD-1/L1 interface, competitive sorting with PD-L1 was performed (Extended Data Fig. 2e). As shown in Extended Data Fig. 2e, 60-fold molar excess unlabeled PD-L1 (864 nM) reduced binding to the target protein.

After five rounds of iterative yeast sorting by FACS, final populations were plated on SD-Trp-Ura agar. Approximately 30 morphologically distinct colonies were randomly isolated for Sanger sequencing. Among these clones, four unique binders were identified and then expressed in *Escherichia coli* for structural analysis and functional profiling (Extended Data Table 1). All designs were soluble and purified by Ni^2+^-NTA chromatography. First, the Rosetta-designed models of the four binder complexes were compared with AF2 predictions (Fig. 2a). Data showed that only one model, PD-L1-5, was inconsistent with AF2. PD-L1-5 exhibited exceptionally high thermal stability (Tm > 95°C) and binding affinity (29.9 nM) (Fig. 2). Binding affinities to PD-L1 assessed by biolayer interferometry were 1.15 pM, 817 pM, 29.9 nM, and 275 nM (Fig. 2b). Consistent with these results, the ultra-high-affinity binder PD-L1-3 was additionally validated by SPR (Extended Data Fig. 3). The circular dichroism spectra of the purified PD-L1 binders matched designed models with melting temperatures (Tm) values exceeding 95°C (Fig. 2c, d).

**Figure 2.**
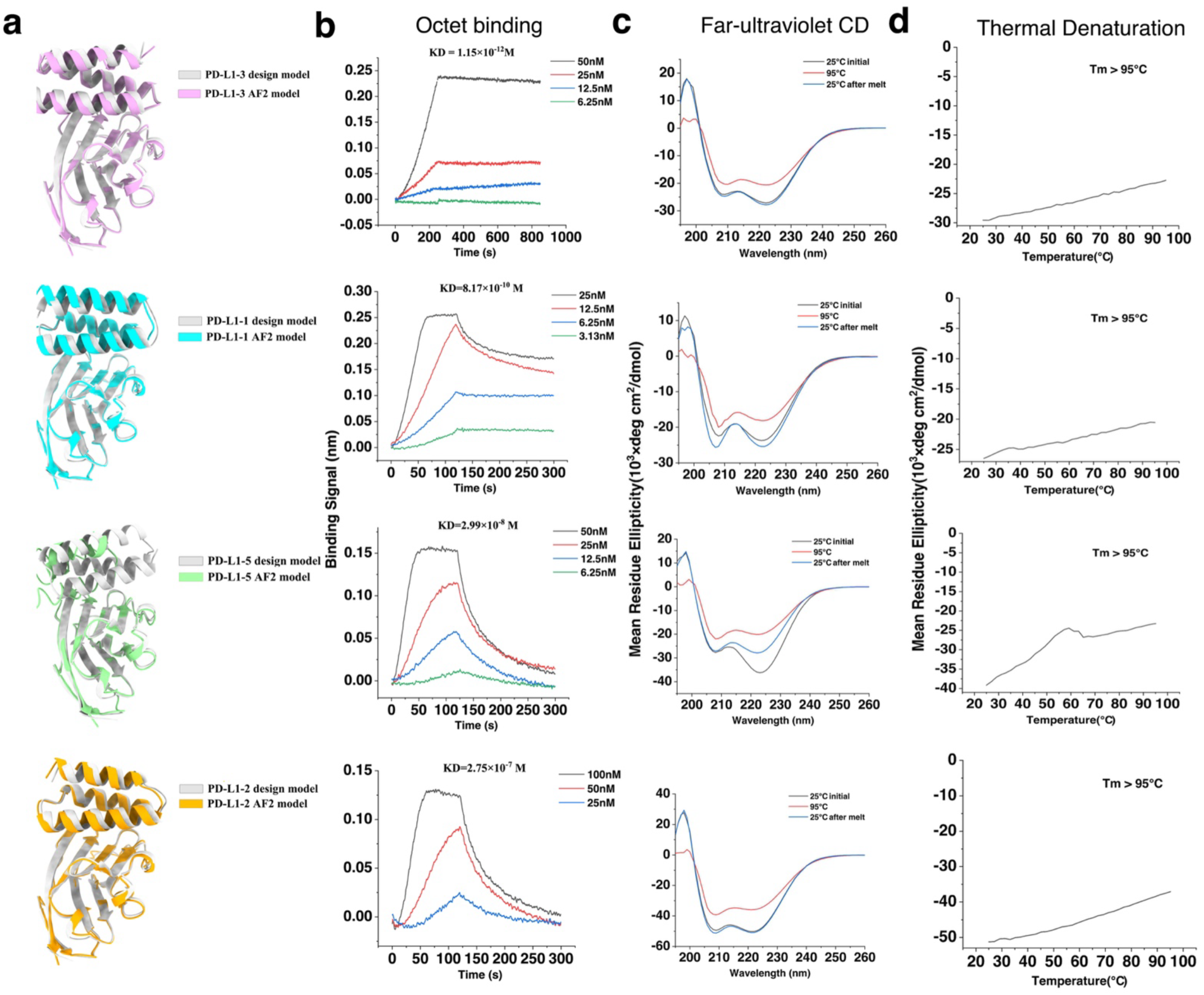
The characterization of de novo designed binders without further experimental optimization. **a,** The de novo designed binders against the interface of PD-1/PD-L1. Grey, Rosetta design model; colors, AF2 prediction model. **b,** Binding of purified miniproteins to the PD-L1 monitored with BLI. **c,** Circular dichroism spectra at different temperatures (left) and CD signal at 222-nm (right), as a function of temperature for the designs. The designs are highly thermostable.

### High-resolution structural validation

High-resolution structure is crucial for evaluating the accuracy of computational protein design. The co-crystal structure of the ultra-high affinity binder PD-L1-3 in complex with the human PD-1 ectodomain was determined using X-ray diffraction (XRD) at 1.77-Å resolution (Fig. 3a and Extended Data Table 2). The co-crystal structure of PD-L1-3 in complex with PD-L1 matched the Rosetta design model with near-atomic accuracy (Cα RMSD = 0.564 Å across the complex; Fig. 3b). PD-L1-3 bound the PD-1/PD-L1 interface through hydrophobic and polar interactions that were recapitulated in computational design models (Fig 3c, d). Although PD-L1-3 binding overlaps with PD-L1’s native receptor site, side-chain interactions differ from native PD-1/PD-L1 complexes. Close agreement between experimental and computational structures indicates that physics-based approaches can design proteins with near-atomic accuracy. This suggests de novo design of proteins with sub-picomolar affinity is achievable without iterative optimization.

**Figure 3.**
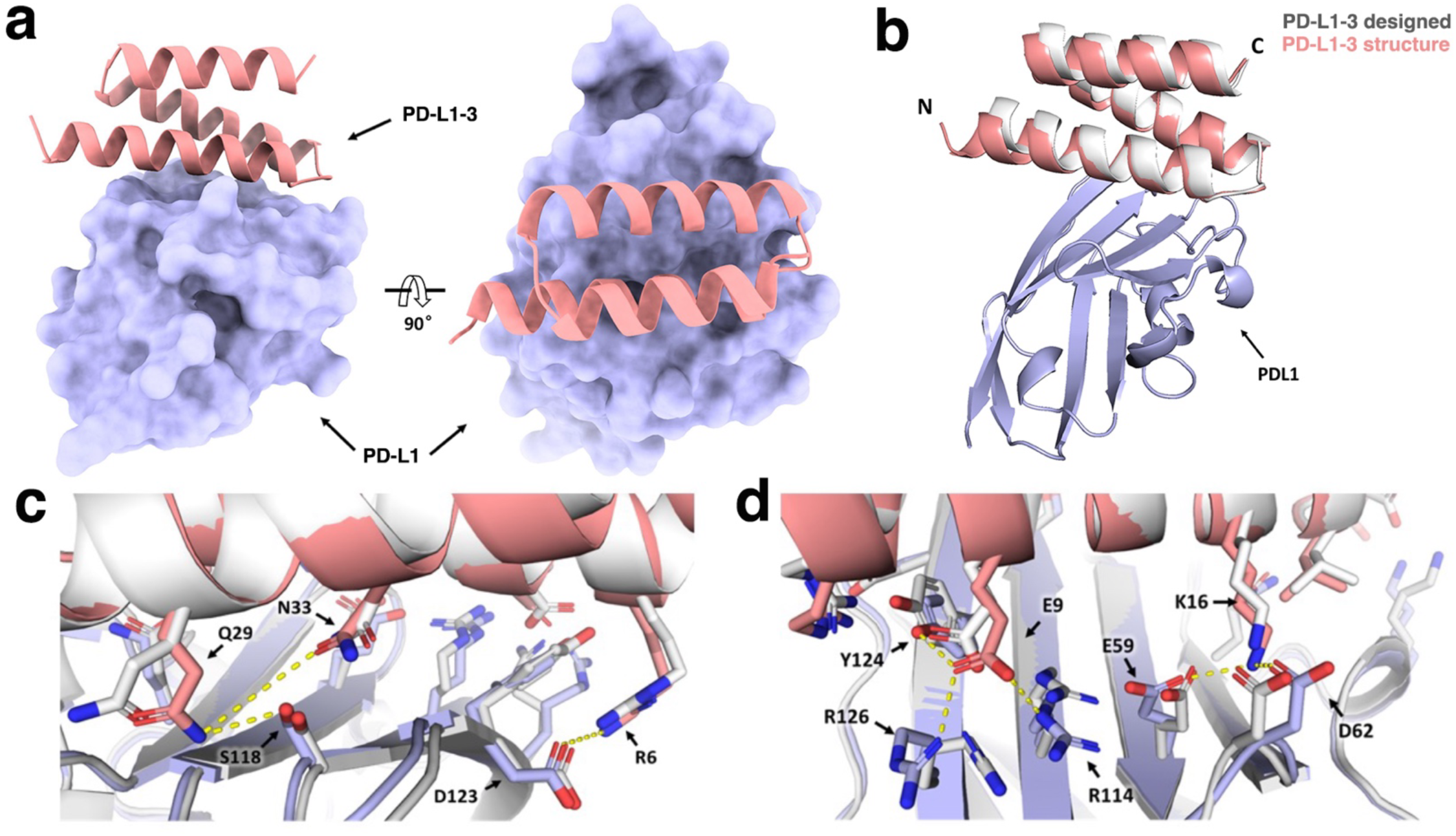
High-resolution structure of miniprotein binder in complex with PD-L1 target protein. **a,** Presentation of solved PD-L1/PD-L1-3 crystal structure: PD-L1 was colored light blue and shown as surface, PD-L1-3 was colored salmon and shown as cartoon. **b,** Superposition of PD-L1/PD-L1-3 complex crystal structure and its design structure, RMSD=0.564. **c-d,** Zoom-in views for the details of the PD-L1/PD-L1-3 interface in crystal structure (light blue for PD-L1 and salmon for PD-L1-3) and design model (gray). Interaction involved in the interface were shown as broken yellow lines, the main chain showed as cartoon and the side chains show as sticks.

### The target specificity, deep tissue penetration, and prolonged tumor retention of the ultrahigh-affinity de novo binder *in vivo*

The de novo-designed binder PD-L1-3, with ultra-high affinity, excellent thermal stability and low molecular weight, was further explored for its dynamic and specific *in vivo* PD-L1 binding by PET-CT. ^68^Ga radiolabeling yielded the PD-L1-specific [^68^Ga]Ga-THP-PD-L1-3 (Fig. 4a and Extended Data Fig. 4), exhibiting specific binding to hPD-L1-positive human lung cancer A549 cells (A549^PD-L1^) versus hPD-L1-negative control (A549) (Fig. 4b). [^68^Ga]Ga-THP-PD-L1-3 achieved saturation binding to PD-L1 on A549^PD-L1^ cells within 30 min, with extended incubation (60-120 min) not increasing PET signal (Fig. 4b). Specificity was validated by competition assays with 1000-fold excess of PD-L1 antibodies Atezolizumab^46^, CS001^47^, APN09^48^, or unlabeled PD-L1-3. Pre-treatment with Atezolizumab, CS001 or PD-L1-3 abolished binding signals (Fig. 4c)

**Figure 4.**
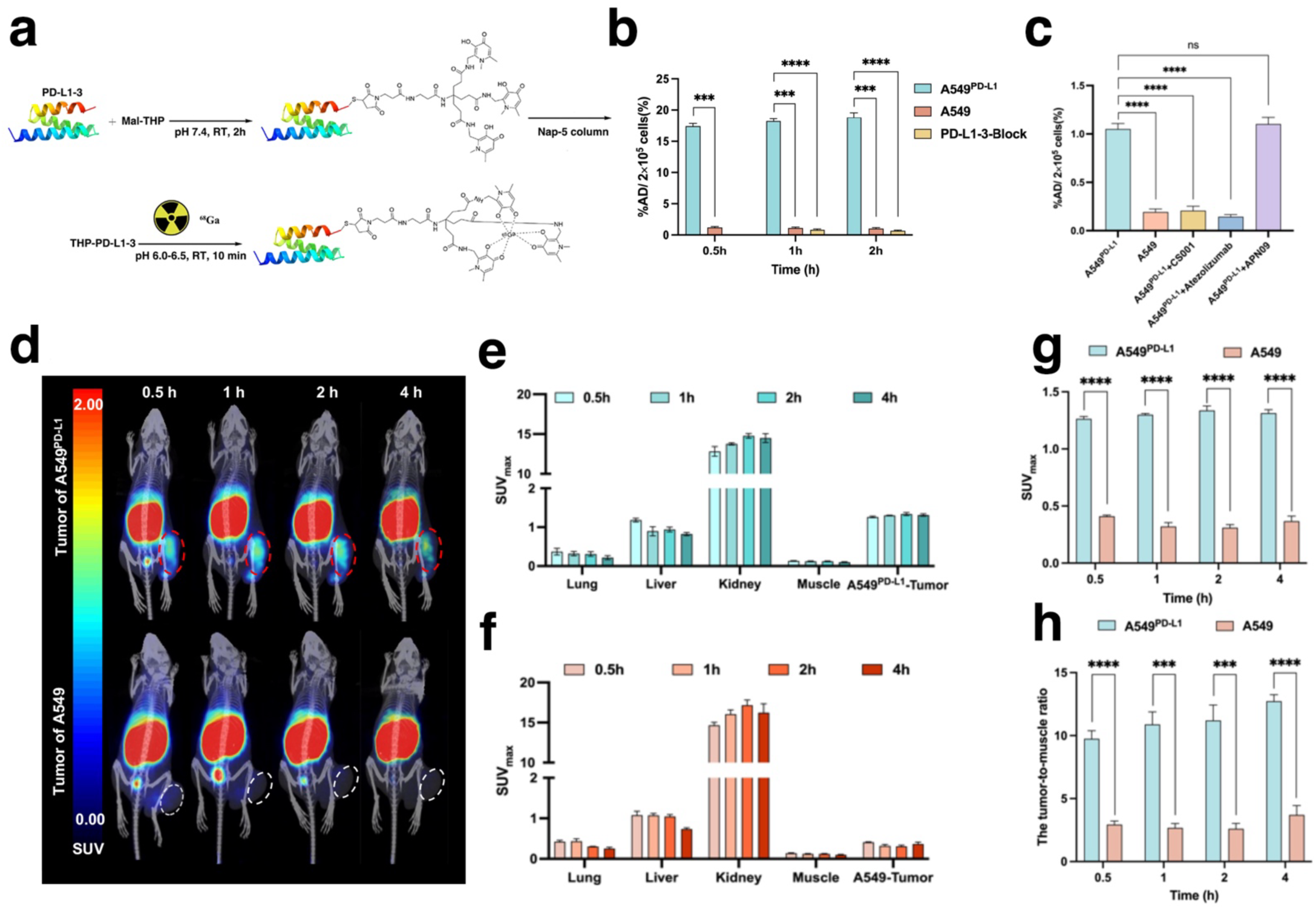
Characterization of [^68^Ga]Ga-THP-PD-L1-3 in vitro and in vivo. **a,** Schematic representation of [^68^Ga]Ga-THP-PD-L1-3 preparation. **b,** Binding specificity of [^68^Ga]Ga-THP-PD-L1-3. n = 4; error bars represent mean ± SD. Comparison between groups was calculated using the two-tailed unpaired Student’s t test. ****P ≤ 0.0001. ***P ≤ 0.001. **c,** Cellular uptake of [^68^Ga]Ga-THP-PD-L1-3 after pre-incubation for 1 h with excess CS001, Atezolizumab, and APN09. n = 4; error bars represent mean ± SD. Statistical comparisons were performed using a two-tailed unpaired Student’s t test. ****P ≤ 0.0001. ns, not significant. **d,** Micro-PET imaging of [^68^Ga]Ga-THP-PD-L1-3 in A549^PD-L1^ (top) and A549 (bottom) at different time points. **e-f,** SUVmax of [^68^Ga]Ga-THP-PD-L1-3 in organs of A549^PD-L1^ and A549 mice at different time points. n = 3, error bars indicate the mean ± SD. **g-h,** Comparison of tumor uptake between A549^PD-L1^ and A549 at different time points and tumor-to-muscle ratios at corresponding time points. n = 3; error bars represent mean ± SD.

At 30 min post-injection, [^68^Ga]Ga-THP-PD-L1-3 exhibited >5-fold enhanced tumor-to-muscle and tumor-to-blood uptake ratios (P < 0.05) with sustained tumor uptake (>4 h) (Fig. 4d). Pronounced renal uptake indicating rapid renal clearance of the unbound tracer, while moderate hepatic uptake resulted from nonspecific gallium-protein binding (Fig. 4d-f). Absence of uptake in PD-L1-negative A549 tumors confirmed *in vivo* specificity, further supported by biodistribution in KM mice and rats (Extended Data Fig 5a, b). Pharmacokinetics revealed biphasic kinetics: α-phase t₁/₂ = 1.016 min, β-phase t₁/₂ = 15.09 min, indicating superior rapid tissue penetration and short circulatory half-life of PD-L1-3 (Extended Data Fig 5c).

Furthermore, elevated tumor uptake and favorable tumor-to-background ratios were observed 30 min post-injection, with sustained in vivo uptake in PD-L1-positive tumors lasting ≥4 h (Fig. 4g, h). Comparisons of tumor uptake between A549^PD-L1^ and A549 at multiple time points revealed slightly increasing tumor-to-muscle ratios of [^68^Ga]Ga-THP-PD-L1-3 over 4 h, demonstrating the binding specificity of the ultra-high-affinity binder PD-L1-3 (Fig. 4h).

To further explore tumor penetration advantages of the ultra-high-affinity binder PD-L1-3, [^68^Ga]Ga-THP-PD-L1-3 uptake was evaluated in large, medium, and small MC38^PD-L1^ tumors (Extended Data Fig. 6). In large MC38^PD-L1^ tumors, elevated radiotracer uptake showed significantly increased tumor-to-muscle ratios with spatial heterogeneity; quantifiable uptake in medium and small tumors confirmed PD-L1-3 penetration capacity (Extended Data Fig. 6). In medium tumors, pre-administration of atezolizumab significantly reduced radiotracer uptake, demonstrating [^68^Ga]Ga-THP-PD-L1-3 binding specificity *in vivo*.

To assess the sensitivity of [^68^Ga]Ga-THP-PD-L1-3 in detecting differential PD-L1 expression, we employed human xenograft models with graded PD-L1 expression (Extended Data Fig. 7a,b). A decreasing uptake gradient was observed: MC38^PD-L1^ > HCC827 > H1975, which correlated with PD-L1 expression levels quantified via Western blot (Extended Data Fig. 5c). Critically, [^68^Ga]Ga-THP-PD-L1-3 detected murine PD-L1-expressing MC38 cells *in vivo*, as quantitatively validated via BLI demonstrating the binding of PD-L1-3 to murine PD-L1 (KD=8.579×10^-10^ M)(Extended Data Fig. 5d). These results confirm its applicability for interrogating native immune microenvironments in murine systems.

The safety and immunogenicity of [^68^Ga]Ga-THP-PD-L1-3 were evaluated (Extended Data Fig. 8 and 9). We monitored longitudinal clinical parameters including routine hematology, histopathology (H&E staining), and quantified anti-PD-L1-3 antibody titers post-administration. PD-L1-3 exhibited favorable safety and low immunogenicity (Extended Data Fig. 8 and 9). The rapid specific uptake and prolonged tumor retention of [^68^Ga)Ga-THP-PD-L1-3 in PD-L1-positive tumors *in vivo* first reveals the advantages of an ultra-high-affinity de novo binder for diagnostic applications, prompting further evaluation of its clinical translation.

### Biodistribution and dynamic binding of de novo binder PD-L1-3 in human malignancies

Here, we report the first-in-human PET/CT study (NCT06383598) of [^68^Ga]Ga-THP-PD-L1-3, confirming the clinical superiority of this ultra-high-affinity de novo binder. Three patients with locally advanced/metastatic LUAC or CSCC were enrolled (April 1-May 31, 2024; Extended Data Table 3). Consistent with prior ^89^Zr-atezolizumab data^49^, [^68^Ga]Ga-THP-PD-L1-3 exhibited minimal physiologic uptake in brain, muscle, lung, and subcutaneous tissue, with progressive tracer accumulation in kidneys, spleen, and liver (Fig. 5a, b). Whole-body (WB) dynamic PET in a CSCC patient achieved visual detection threshold at 300 s post-injection. The most striking finding observed was the cumulative tumor uptake of [^68^Ga]Ga-THP-PD-L1-3 exhibited a persistently elevated contrast from 5 minutes to 3 hours (Fig. 5c, Extended Data Fig. 10 and Supplementary Video). In contrast, ^89^Zr-atezolizumab achieved peak tumor-to-background ratio on day 7^49^. Furthermore, even the cyclic peptide-based PD-L1 tracers such as WL-12^50,51^, DK223^52^ and its derivative NK224^53^ with lower molecular weight compared to PD-L1-3, their blood clearance, maximum radiotracer accumulation in tumors and persistent tumor retention were not comparable to the de novo binder. These data further revealed the advantages of the ultra-high-affinity de novo binder in tissue permeability and sensitivity for PD-L1 recognition in the body.

**Figure 5.**
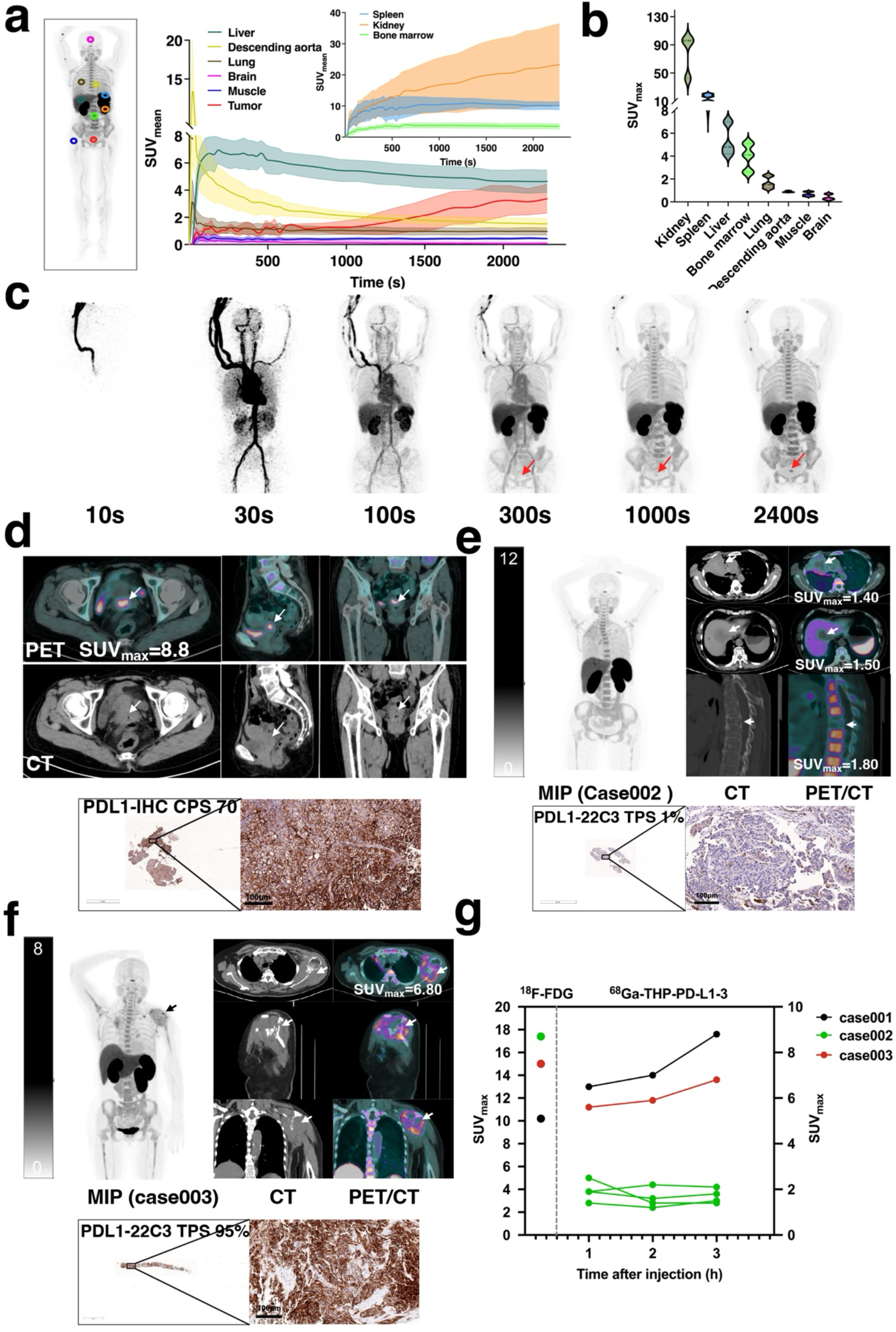
[^68^Ga]Ga-THP-PD-L1-3 biodistribution and tumor uptake. **a,** SUVmean (95% CI) in different organs at 0-40 min post-injection in a CSCC patient. **b,** Uptake of [^68^Ga]Ga-THP-PD-L1-3 in different organs as indicated by SUVmax at 3 h (n=3). **c,** Dynamic PET/CT scan of [^68^Ga]Ga-THP-PD-L1-3 in the CSCC patient over 0-40 min (lesion indicated by red arrow). **d,** Images from a woman with CSCC (left) and a PD-L1 CPS of 70 (right; scale bars, 100µm; IHC was performed once). SUVmax of primary tumor was 8.8 (white arrow). **e,** Images from a woman with LUAC (top) and a PD-L1 TPS of 1% (bottom; scale bars, 100µm; IHC was performed once). SUVmax values: primary tumor in the right lung was 1.4 (white arrow); metastatic tumors in liver and bone were 1.5 and 1.8 (white arrows). **f,** Images from a woman with LUAC (top) and a PD-L1 TPS of 95% (bottom; scale bars, 100µm; IHC was performed once). SUVmax of metastatic tumor in the left scapula was 6.8 (white arrow). **g,** Representative PET/CT images in PD-L1-positive patients 3 h post-injection show increasing tumor uptake over time.

Remarkable tumor uptake of [^68^Ga]Ga-THP-PD-L1-3 (SUVmax: 8.8) was observed 3 h post-injection in a high PD-L1 CSCC patient (CPS=70, IHC-comfirmed; Fig. 5d). The LUAC patient with high PD-L1 expression exhibited a higher tumor lesion uptake of [^68^Ga]Ga-THP-PD-L1-3 (TPS: 95%, SUVmax: 6.8) than the patient with lower PD-L1 expression (Fig. 5e, f). The primary tumor and metastases in the liver and bone in the LUAC patient with low PD-L1 TPS grades, showed low uptake of the radioactive tracer (TPS: 1%; SUVmax: 1.4, 1.5, and 1.8) (Fig. 5e, g). Assessment of the tumor-to-heart, tumor-to-muscle and tumor-to-background ratios revealed that [^68^Ga]Ga-THP-PD-L1-3 exhibited excellent uptake ratios and cumulative tumor uptake in the patients from 1 to 3 hours post-injection (Extended Data Fig. 10). These clinical data showed strong correlation between tracer uptake and PD-L1 expression (Fig. 5g and Extended Data Fig. 10), validating the real-world specificity of this ultra-high-affinity de novo binder. Moreover, [^68^Ga]Ga-THP-PD-L1-3 demonstrated favorable safety profiles, as evidenced by the absence of related adverse events or clinically detectable pharmacological effects in any patients (Extended Data Table 4). No significant changes were observed in vital signs, laboratory tests, or electrocardiogram.

## Conclusions

Our success in designing near-subpicomolar affinity anti-PD-L1 protein binder indicates that physics-based methods, with no need for iterative experimental optimization, can accurately design monomeric proteins with exceptionally high binding affinity. Previously, Cao et al. reported a significant study using a physics-based approach to achieve the goal of direct computational design of high-affinity binders against the spike receptor-binding domain of SARS-COV-2, with affinities ranging from 100 nM to 2 µM, starting from the target structural information alone. Following further optimization through SSM and combinatorial library screenings, the binders with affinities ranging from 100 pM to 10 nM were obtained^28^. Moreover, deep-learning methods such as RF Hallucination^26,31^ and RFdiffusion^5^, with improved experimental success rates, can directly design de novo protein binders with affinities ranging from 27 nM to 1.4 µM. Very recently, using fluorescence polarisation binding assays, Susana and her colleagues showed that RFdiffusion can directly design picomolar affinity binders (Kd < 500pM) for helical peptides^30^. By combining a physics-based de novo binder design method with physics and DL-based metric filters, our work represents a major step forward towards the goal of directly designing near sub-picomolar protein binders using computational methods, starting from the target structural information alone. To our knowledge, the PD-L1-binding protein PD-L1-3 is the highest-affinity protein binder to any target (protein, peptide, or small molecule) achieved directly by computational design, with no need for iterative experimental optimization. More importantly, the physics-based de novo binder design method and the high-affinity binders can provide a starting point for investigating the fundamental physical chemistry of protein-protein interactions of these exceptionally high-affinity binders, as well as for developing and assessing computational models of protein-protein interactions.

Over the past three decades, antibodies have risen to prominence, delivering dramatic breakthroughs for the treatment of various human diseases and gaining importance as a key therapeutic modality for many drug developers^54^. The de novo designed binder in this work has shown potential advantages over antibodies, such as higher binding affinity, thermostability, tumor permeability, and simplified, rapid, and economical expression in *E. coli*. It can be regarded as ideal tools, functioning as signaling pathway antagonists in their monomeric protein format and as tunable agonists in their multimeric formats, and they can be used in diagnostics and therapeutics for indications including oncology, autoimmunity, chronic inflammatory diseases, and infectious diseases. Our preliminary preclinical and clinical data of immunoPET imaging with [^68^Ga]Ga-THP-PD-L1-3 further validates the benefits of the de novo designed binder with exceptionally high binding affinity. Although preclinical and clinical data of PD-L1 tracers, such as cyclic peptide-based tracers WL-12^50,51^ or DK223^52^ and its derivative NK224^53^, and antibody-based tracer Atezolizumab^49,55,56^ have been previously reported, none of these tracers has been approved by the FDA. Comparison of these results showed that [^89^Zr]-Atezolizumab exhibited excellent binding specificity and persistent tumor retention, but the maximal systemic clearance of [^89^Zr]Zr-atezolizumab was reached at 6 ± 1 days post-injection^49,55,56^. Owing to their lower molecular weight relative to antibodies, the cyclic peptide-based PD-L1 tracers exhibited faster blood radiotracer clearance (∼90% clearance within 60 min) and quicker maximum radiotracer accumulation in tumors (at 60 min)^52^. However, the binding affinity and the capacity of persistent tumor retention were significantly lower to those of antibodies. DK223 exhibited 1 nM binding affinity. Although DK223 maintained relatively high tumor retention even at 180 min, the radiotracer accumulation in tumors decreased at 90 min^52^. Compared with these PD-L1 tracers, the ultra-high-affinity de novo binder PD-L1-3 composed of 79 amino acids exhibited superior binding affinity (1.15 pM). Our preclinical and clinical data demonstrated that the PD-L1-3 exhibited excellent binding specificity, tumor penetration and persistent tumor retention, reaching maximum tumor accumulation at 30 min post-injection and showing progressively enhanced tumor retention for 4 h. The most striking finding in this study is that, although PD-L1-3 has a >5-fold greater molecular weight (9.43 kDa) compared with peptide tracers (DK221:2 kDa; WL12:1.8 kDa), it exhibited unprecedented clearance kinetics, with fast-phase (t₁/₂=1.016 min) and slow-phase (t₁/₂=15.09 min) half-lives. These findings have been further supported by our clinical data that PET images of the CSCC patient revealed high radiotracer accumulation in tumors as early as 5 min after injection, with continuous increases in target-positive tumor radiotracer accumulation, tumor-to-heart ratio, tumor-to-muscle ratio, and tumor-to-background ratio over a 3-hour period.

Our data revealed three core advantages of ultra-high-affinity de novo binder: precise binding specificity, enhanced tissue/tumor penetration, and prolonged target-positive tumor retention across preclinical models and clinical applications. More generally, the ability to directly design ultrahigh-affinity binders to target proteins confers transformative potential across multiple disciplines of biotechnology and medicine that reliant on affinity-driven molecular recognition, providing a foundational framework for the next generation of diagnostics and therapeutics transcending natural evolutionary constraints.

## Methods

### De novo binder design

The crystal structure (PDB:4Z18) of human PD-L1 was refined with the Rosetta FastRelax protocol with coordinate constraints. The targeting chain was extracted and used as the starting point for docking and design. To run PatchDock, the de novo scaffold library of 1739 proteins (with residue lengths ranging from 56 to 65), generated as previously described^33,34^, was first mutated to poly-valine, and default parameters were then used to generate the raw docks. These docks served as the seeds for the initial positioning of the scaffolds during RifDock. The RIF was generated by RifDock, during which billions of individual disembodied amino acids were docked against the PD-1 binding region of PD-L1 that we had selected. If the amino acids passed a specific energy cutoff value (-1.5 Rosettta energy unit), the corresponding inverse rotamers were generated and their backbone coordinates stored in a six-dimensional spatial hash table for rapid look-up. The protein scaffold library was docked into the field of inverse rotamers using a branch-and-bound algorithm, transitioning from low-resolution to high-resolution spatial grids. These docked conformations were further optimized using the Rosetta FastDesign protocol to generate shape and chemically complementary interfaces, activating between side-chain rotamer optimization and gradient-descent-based energy minimization.

In the sequence design protocol, an alternative Rosetta FastDesign protocol with several improvements, including an enhanced repulsive energy ramping strategy^57^, upweighting cross-interface energies, a pseudo-energy term penalizing buried unsatisfied polar atoms^58^, and a sequence profile constraint based on native protein fragments^59^, was performed to further optimize the binders. Computational physic-based metrics of the design models were calculated using Rosetta^44^, which includes ddG, shape complementary and interface buried SASA, score_per_res, contact molecular surface, contact_patch, interface_buried_sasa, interface_sc, ss_sc, mismatch_probability, worst9mer and worst9mer_helix for design selection.

To further optimize the designed binders and expand exploration, an intensified search around the best-designed interfaces was performed. The binding energy and interface metrics for all the continuous secondary structure motifs of the designed binders were calculated. Using the target structure as the reference, motifs that interact well with the target were extracted and clustered using an energy-based TMalign-like clustering algorithm^60^, with a similarity score threshold of 0.7. The 1188 best motifs were selected, and the scaffold library was superimposed onto these motifs by using MotifGraft mover^61^. The Motif-Grafted binders were further optimized by the alternative Rosetta FastDesign protocol as mentioned above. All the designed binders were evaluated using physics-based and DL-based metrics, specifically Rosetta^44^ and DAN-Bert^42^. Based on the above metrics, the top 100,000 designs were selected and ordered for DNA synthesis from Agilent Technologies.

### DNA library preparation

To avoid the biased amplification of short DNA fragments during PCR reactions, all protein sequences were padded to a uniform length (65 amino acids) by adding a (GGGS)n linker at both N and C terminals of the designs. The protein sequences were reversed translated and optimized using DNAworks2.0^62^ with the S. cerevisiae codon frequency table. The oligo libraries were amplified using Kapa HiFi Polymerase (Kapa Biosystems) with a qPCR machine (ABI 7500). In detail, the libraries were first amplified in a 25 µl reaction, and the PCR reaction was terminated when the reaction reached half of the maximum yield to avoid overamplification. The PCR product was loaded onto a DNA agarose gel. The band with the expected size was cut out and DNA fragments were extracted using QIAquick kits (Qiagen). The DNA product was then re-amplified as before to generate enough DNA for yeast transformation. The final PCR product was cleaned up with a QIAquick Clean up kit (Qiagen). For the yeast transformation, 2-3 μg of linearized modified pETcon vector (pETcon3) and 6 μg of purified PCR product were transformed into EBY100 yeast strain using the protocol as previously described^25,28,63^.

### Yeast surface display

*S. cerevisiae* EBY100 strain cultures were grown in C-Trp-Ura media supplemented with 2% (w/v) glucose. To induce the designed binder expression on yeast cell surface, yeast cells were centrifuged at 6,000g for 1 min and resuspended in SGCAA medium supplemented with 0.2% (w/v) glucose at the cell density of 1 × 10^7^ cells per ml and induced at 30 °C for 16-24 h. Cells were washed with PBSF (PBS with 1% BSA) and firstly incubated with biotinylated PD-L1 (Acro Biosystems). After being washed, cells were subsequently incubated together with anti-c-Myc fluorescein isothiocyanate (FITC, Abcam) and avidin-phycoerythrin (PE, ThermoFisher) at the dilution ratio of 1:100. The yeast cells were sorted twice with a high concentration (1.44µM) of biotin-labelled PD-L1 target protein, followed by two additional sortings at different concentrations of the PD-L1 target protein. After the cells were enriched with 144 pM of PD-L1 target protein in the fifth sorting, competition sorting was performed. The biotinylated PD-L1 target protein (14.4 nM) was first incubated with excess amounts of PD-1 competitor atezolizumab in the range from 864 nM to 8.64 nM for 10 min. Then, the mixture was incubated with washed yeast cells for labeling. The final sorting pools were plated on C-trp-ura plates, and the sequences of individual clones were determined by Sanger sequencing.

### Protein expression

Genes encoding the designed protein sequences were cloned into pET-29b(+) E. coli plasmid expression vectors (GenScript). For all the designed proteins, the sequence of the C-terminal tag used is SGLEHHHHHH. Plasmids were then transformed into chemically competent E. coli Lemo21 cells. The *E. coli* cells were grown in LB medium at 37 °C until the cell density reached 0.6 at OD600. Then, IPTG was added to a final concentration of 1 mM and the cells were grown overnight at 22 °C for expression. The cells were harvested by spinning at 4,000 × g for 10 min and then resuspended in BugBuster® Protein Extraction Reagent (Merck) with DNase (Millipore) and protease inhibitor tablets (Merck). The cells were lysed with a Qsonica Sonicators sonicator for a total of 4 minutes (2 minutes on time, 10 sec on, 10 sec off) with an amplitude of 80%. The soluble fraction was then clarified by centrifugation at 20,000g for 30 minutes. The soluble fraction was purified by immobilized metal affinity chromatography (ThermoFisher), followed by FPLC size-exclusion chromatography (Superdex 75 10/300 GL, Cytia). All protein samples with the purity higher than 95% were characterized by SDS-PAGE. Protein concentrations were determined by absorbance at 280 nm, measured using a NanoPhotometer (Implen), and predicted using extinction coefficients.

### Biolayer interferometry

Biolayer interferometry binding data were collected using an Octet R8 (Sartorius) and processed using the instrument’s integrated software. The biotinylated PD-L1 (Acro Biosystems) was loaded onto streptavidin-coated biosensors (SA Sartorius) at 2µg/ml in binding buffer (10 mM HEPES (pH 7.4), 150 mM NaCl, 3 mM EDTA, 0.05% surfactant P20, 0.5% non-fat dry milk) for 600s. The binders were diluted from concentrated stocks into the binding buffer. After baseline measurement in the binding buffer alone, the binding kinetics were monitored by dipping the biosensors loaded with the PD-L1 target protein into wells at the indicated concentration (association step) and then dipping the sensors back into baseline/buffer (dissociation step). Data were analysed and processed using Octet Analysis Studio 13.0.1.35.

### Affinity measurements by SPR

SPR analyses were performed with a Biacore 8K instrument (Cytiva), where data were processed using its integrated evaluation software. Biotinylated recombinant human PD-L1(AcroBiosystems, PD1-H5229) was immobilized on a CM5 sensor chip (Cytiva). Serially diluted protein binders were injected over the sensor surface in 1× HBS-EP^+^ buffer (Cytiva BR100669).

### Circular dichroism

Far-ultraviolet circular dichroism measurements were carried out using a JASCO-1500 equipped with a temperature-controlled multi-cell holder. Wavelength scans were measured from 260 to 190 nm at 25, 95°C and again at 25°C after fast refolding (∼5 min). The dichroism signal at 222 nm was monitored during temperature melts in steps of 2°C/min with 30s of equilibration time. Wavelength scans and temperature melts were performed using 0.3 mg/ml protein in PBS buffer (20mM NaPO4, 150mM NaCl, pH 7.4), with a 1 mm path-length cuvette. Melting temperatures were determined by fitting the data with a sigmoid curve equation. All of the designs retained more than half of the mean residue ellipticity values, which indicates that the Tm values are greater than 95°C.

### ELISA

To assess binder immunogenicity, C-terminal biotinylated PD-L1-3 was immobilized on streptavidin-coated plates (Thermo Fisher #15124) at 2.5 µg/mL in a total volume of 100 µL per well and incubated at 4℃ overnight. Plates were washed with wash buffer (TBS + 0.1% (w/v) BSA + 0.05% (v/v) Tween20) and blocked with 200 µL/well blocking buffer (TBS + 2% (w/v) BSA + 0.05% (v/v) Tween20) for 1 h at room temperature. Plates were rinsed with wash buffer using 200 µL/well, and 100 µL of 1:100 diluted sera samples in blocking buffer were added to respective wells. For a positive control, Atezolizumab was serially diluted 1:5 starting at 200 ng/mL in 100 µL of blocking buffer. All samples were incubated for 1 hour at room temperature. Plates were washed using 200 µL of wash buffer per well. For the serum samples, HRP-conjugated goat anti-mouse IgG antibody (Thermo Fisher #SA5-10276) was diluted 1:500 in blocking buffer, and 100 µL was incubated in each well at room temperature for 30 minutes. For the positive control, HRP-conjugated goat anti-human IgG antibody (Thermo Fisher # SA5-10273) was diluted 1:500 in blocking buffer, and 100 µL was incubated in each well at room temperature for 30 minutes. Plates were rinsed with wash buffer, and 100 µL of TMB (Thermo Fisher # N301) was added to each well for 30 min. The reaction was quenched by adding 100 µL of 0.16M H2SO4. Optical densities were determined at 450 nm on an Infinite M200 Pro plate reader (Tecan Instruments).

### Crystallization and structure determination of the PD-L1-3 in complex with PD-L1

The crystallization experiments were performed using the hanging-drop vapour diffusion method at 18 °C with 2-μl drops containing 1 μl protein solution (6.5 mg/ml) and 1 μl reservoir solution equilibrated over 100 μl reservoir solution. Initial crystallization hits of PD-L1 with PD-L1-3 were found from the Morpheus Kit (Molecular Dimensions) B4. Qualified crystals of PD-L1 with binder were obtained in the reservoir buffer containing 0.03M Sodium fluoride, 0.03M Sodium bromide, 0.03M Sodium iodide, 0.1 M (Imidazole/MES) pH6.5, 12.5% v/v MPD, 12.5% PEG1000, 12.5% w/v PEG 3350 within 2 weeks. For data collection, the crystals were soaked in a cryoprotectant solution containing the reservoir buffer supplemented with 20% Glycerol before flash-freezing with liquid nitrogen. Diffraction data were collected at the Shanghai Synchrotron Radiation Facility (Shanghai, China) beamline BL02U1 at a wavelength 0.97915 Å, and processed with the XDS^64^ and Aimless^65^. The predicted structure of PD-L1 and nanobody by AlphaFold Ⅱ was used as a starting model for the molecular replacement^66^, using Phaser. The structure was refined by iterative cycles of manual model building and refinement in Coot^67^ and Phenix refine^68^. Ramachandran statistics were generated using MolProbit^69^. All structural figures were generated in PyMOL (http://www.pymol.org).

### Preparation and radiolabeling of precursors

The protein solvent was 0.01 M PBS, pre-treated with Chelex 100, and the protein concentration was 2-4 mg/mL. The preparation process of the precursor is depicted in Fig. 4a. The ^68^Ga-labeled precursor can be efficiently synthesized by reacting a free Cys residue at the C-terminal of PD-L1-3 with the maleimide group present in the chelating agent THP-Mal. The protein was added to Mal-THP at a molar ratio of 1:10, with an approximate quantity of 2 mg, and then incubated at 37℃ for one hour. Subsequently, the desired precursor was purified using PD-10 or Nap-5 gel column chromatography. The collected product was then packed in quantities of 100 μg and stored at -80℃ until further use.

The labeling procedure is also depicted in Fig. 4. ^68^GaCl3 (3 mL, 0.05 M HCl) was eluted from a ^68^Ge-^68^Ga generator (1.85 GBq, ITG Co., Ltd, Germany), with the first 1 mL being discarded. Subsequently, 1 mL of gallium eluent (∼37 MBq) was transferred into a 4 mL reaction tube, followed by the addition of 300 μL of 2M NaAc and 100 μg of precursor. The mixture was thoroughly blended and allowed to react at room temperature for 10 minutes. After the completion of the reaction, purification was performed using a PD-10 column. The resulting product passed through a 0.22 µm sterile filter membrane into a sterile vacuum bottle and diluted with saline for administration. Radiochemical purity was measured by radio-thin layer chromatography (radio-TLC).

### Cell binding experiment

A549 and A549^PD-L1^ cell lines were cultured in RPMI-1640 medium with 10% fetal bovine serum and 1% penicillin-streptomycin solution, in an incubator at 37 °C and 5% CO2. Before the cell binding experiment, cells were seeded in 24-well plates (2×10^5^ cells/well) and cultured overnight. After discarding the medium, 0.5 mL of [^68^Ga]Ga-THP-PD-L1-3 solution (74 KBq) was added per well, and the cells were incubated at room temperature. At 30 min, 60 min, and 120 min, the liquid in the well was discarded, and the wells were washed three times with PBS to remove unbound tracer. 200 μL of NaOH solution (0.5 M) was added, the lysate was then transferred to a counter tube, and the radioactivity was measured using a γ-counter. In the blocking group, a sufficient amount of PD-L1-3 and [^68^Ga]Ga-THP-PD-L1-3 solution (74 KBq) were added. In another group, after medium removal, the wells were washed three times with PBS. Antibodies (atezolizumab, CS001, or APN09; n=4/group) at 1000-fold molar excess were added, followed by 0.5 mL of [^68^Ga]Ga-THP-PD-L1-3 solution (74 kBq). After 60-min incubation at room temperature, supernatant was discarded and wells washed three times with PBS. Cell lysis used 200 μL 0.5 mol/L NaOH, radioactivity quantified via γ-counter (model Wizard²®, PerkinElmer) to calculate cellular uptake.

### Micro-PET/CT imaging in vivo

Female BalB/c Nude mice (4-6 weeks old) were ordered from Beijing Vital River Animal Technology (Beijing, China). Animal experiments were conducted in accordance with the regulations of the Animal Care and Use Committee of Peking University Cancer Hospital. A549 and A549^PD-L1^ cells (2×10^6^, 100 *μ*L) were injected into the root of the thigh muscle, respectively. Imaging was performed when the tumor size reached 200-500 mm^3^. Mice were injected with [^68^Ga]Ga-THP-PD-L1-3 (3.7-7.4 MBq, 200 *μ*L) through the tail vein and underwent Micro-PET imaging at 0.5 h, 1 h, 2 h, and 4 h. The mice were initially anaesthetized with 3% isoflurane and then continuously anaesthetized with 1% isoflurane on the imaging bed during imaging. We defined tumor sizes as follows: large tumors (500–600 mm^3^), medium tumors (150–200 mm^3^), and small tumors (50–100 mm^3^). Tumor models were established using MC38^PD-L1^ cells, with atezolizumab (blocking antibody) and MC38 cells as the negative control. Tumor-bearing mice in experimental groups received 7.4 MBq of [^68^Ga]Ga-THP-PD-L1-3 intravenously (i.v.). In the blocking group, 2 mg atezolizumab was co-administered with the tracer. Micro-PET imaging was performed at 0.5, 1, and 2 h post-injection. PET images were acquired for 10 minutes and reconstructed with attenuation correction based on CT data (CT-AC Reconstruction). Regions of interest (ROIs) were drawn on the CT images and then further mapped on PET.

### Clinical trial approval and inclusion criteria of patients

The clinical translational study was approved by the Medical Ethics Committee of Peking University Cancer Hospital (2022KT74-ZY01) and registered at ClinicalTrials.gov (NCT06383598, Date of registration: April 22, 2024). The study was conducted following the guidelines of the Declaration of Helsinki. Written informed consent was obtained from all participants prior to [^68^Ga]Ga-THP-PD-L1-3 PET/CT imaging. The inclusion criteria for volunteers are: (1) age between 18 and 70 years; (2) Eastern Cooperative Oncology Group performance score of 0-1; and (3) patients with solid tumors confirmed by clinical consultation. The exclusion criteria are: (1) severe impairment of liver and kidney function; (2) pregnancy or current lactation; and (3) other unsuitable conditions deemed by researchers. Three cancer patients were enrolled in this study.

### [^68^Ga]Ga-THP-PD-L1-3 prepared for PET/CT imaging in patients

[^68^Ga]Ga-THP-PD-L1-3 was prepared in a GLP environment dispensing hot cell (NMC Ga-68, Tema Sinergie S.p.A, Italy). Patients were injected with the radiotracer (1.52-2.72 MBq/kg) intravenously. Quality control studies were conducted and details can be found in the Supplemental information (Extended Data Fig. 11).

### PET/CT imaging in patients

PET/CT imaging with a 194-cm axial field of view (FOV) was conducted using a total-body PET/CT (uEXPLORER, United Imaging Healthcare, Shanghai, China). After the intravenous injection of [^68^Ga]Ga-THP-PD-L1-3, one patient underwent a 40-minute dynamic PET/CT scan, while all patients underwent subsequent static PET/CT imaging at 2 and 3 hours post-injection, with each scan lasting for 5 minutes. Before the injection, low-dose CT imaging was performed with the following parameters: 50 mAs, 140 kVp, tube current modulation on, and an effective dose of approximately 10 mSv. Additionally, [^18^F]F-FDG PET/CT was performed within one week (with blood glucose levels below 160 mg/dL before injection, and scanned at 60 minutes post-injection).

### First-in-human evaluation of PET/CT imaging in patients

Data processing was conducted by using vendor-provided software MultiModality Workplace (United Imaging, China). The biodistribution of [^68^Ga]Ga-THP-PD-L1-3 was analyzed by drawing the ROIs of major organs and tumor lesions. To analyze and compare data from the images, the maximum single-voxel standardized uptake value (SUVmax) was quantified by drawing ROIs in Vendor-provided software. Attenuation correction for [^68^Ga]Ga-THP-PD-L1-3 radioactive activities was conducted to calculate the organ biodistribution.

## Acknowledgements

We thank Prof. David Baker at the Institute for Protein Design, University of Washington, for providing the visiting scholar program and support in de novo protein design, and Brian Coventry for support in protein design method development. This work was supported by the National Natural Science Foundation of China (8227130865 to L.Z., 82171973 to H.Z.), Capital’s Funds for Health Improvement and Research (2022-2Z-2155 to H.Z.), the Fundamental Research Funds for the Central Universities (030-63233042 to Y.G.).

## Author Contributions

L.Z., Y.H., H.Z., Y.G. conceived the project. L.Z. developed the computational methods. L.Z., F.Z., G.Z., R.Y. designed, screened and experimentally characterized the binders. F.Z., G.Z., R.Y., G.G., X.L., G.Y., T.L., H.L., S.C. performed gene construction, protein expression, purification and characterization. Y.G., Z.S. performed the collection of X-ray data, as well as structure determination. H.Z., F.W. were responsible for the recruitment of patients and image analysis. S.H., J.T., Q.W. were involved in the preparation of radiopharmaceuticals and took part in some animal experiments. B.S., Y.Y. provided research support and supervision. J.F. provided research strategy and funding acquisition support. All authors reviewed and accepted the manuscript.

## Data Availability

The atomic coordinates and experimental data of PD-L1-3 in complex with human PD-L1 has been deposited in the RCSB PDB with the accession numbers 8ZNL.

## Code availability

The Rosetta macromolecular modelling suite (https://www.rosettacommons.org) is freely available to academic and non-commercial users. Commercial licences for the suite are available through the University of Washington Technology Transfer Office. The source code for RIF docking implementation is freely available at https://github.com/rifdock/rifdock. The design scripts and main PDB models, computational protocol, the entire miniprotein scaffold library and all the design models used in this paper can be downloaded from Science Data Bank: https://www.scidb.cn/en/s/2iAZji.

## Supplementary Information

Supplementary material including description of experimental methods is available.

## Extended Data

**Extended Data Figure 1.**
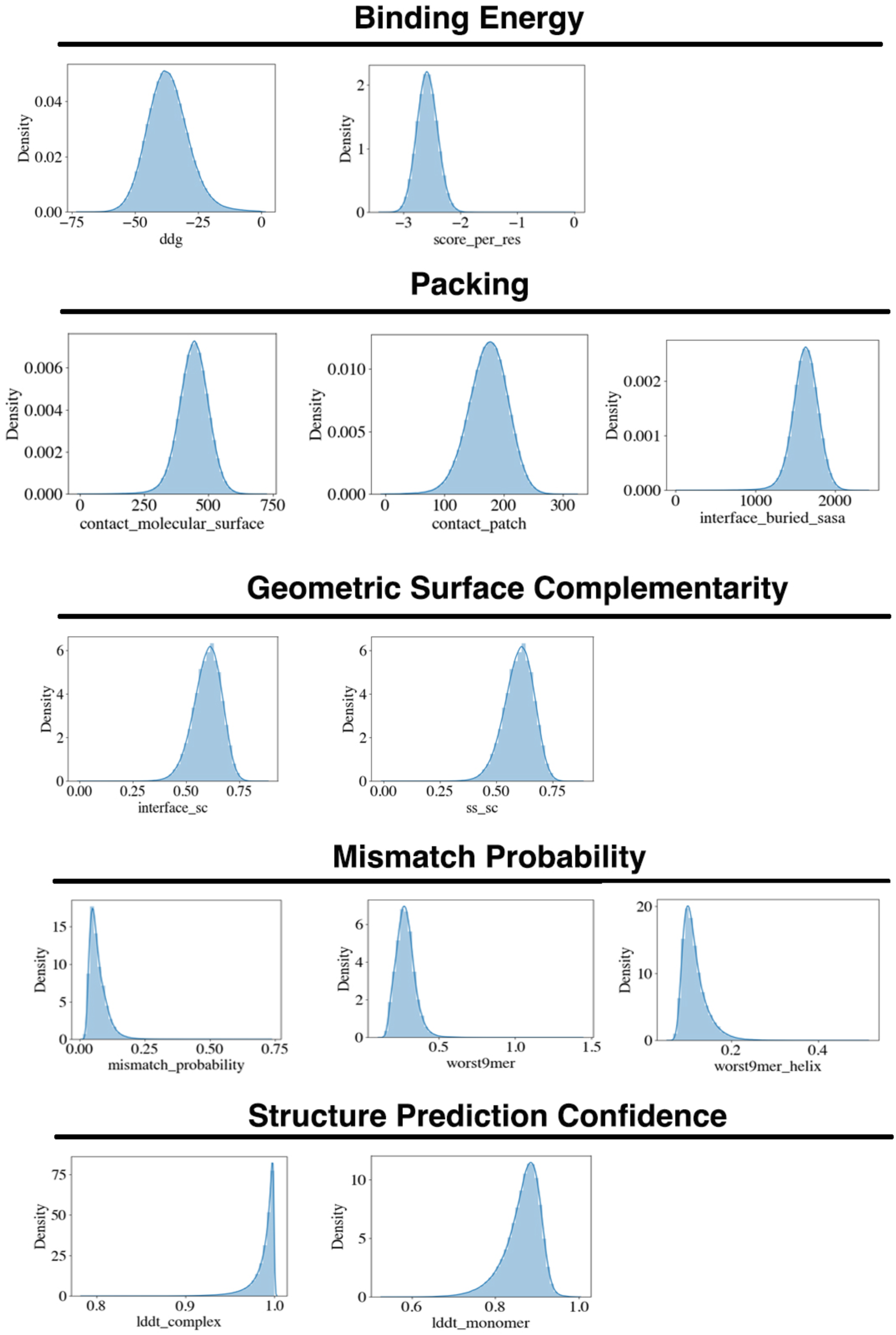
The density plots show the distributions of designed binders. For these de novo designed PD-L1 binders, the distributions of binding energy, packing, geometric surface complementarity, mismatch probability, and structure prediction confidence were calculated based on physics-based and DL-based metrics.

**Extended Data Figure 2.**
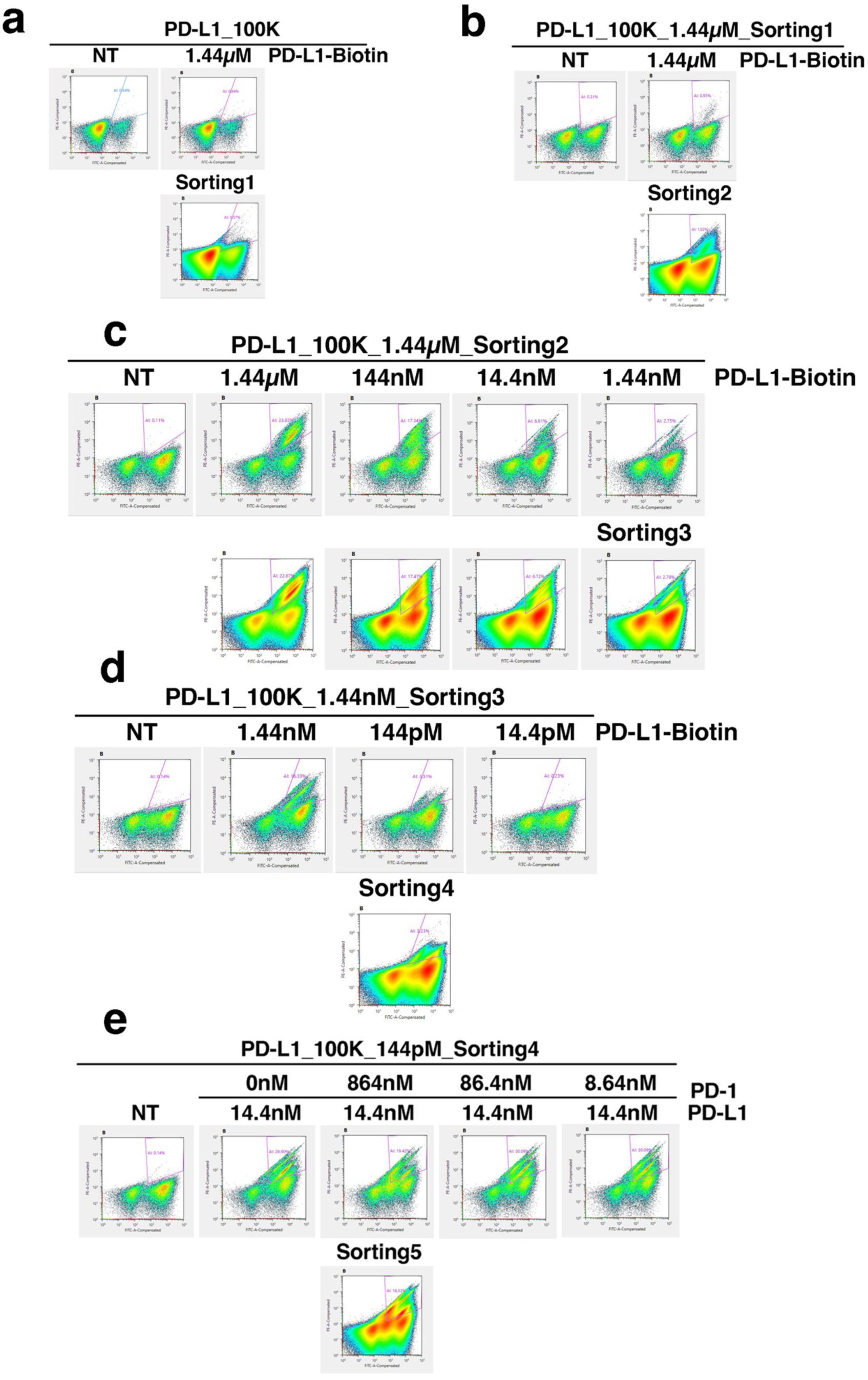
Yeast surface display screening of PD-L1 de novo designed binders. Yeast cells displaying the indicated design were incubated with biotin labeled PD-L1, and PD-L1 binding to cells (Y axis) was monitored by flow cytometry. Approximate sort gates are shown for Sort 1 (**a,** 1.44 µM PD-L1), Sort 2 (**b,** 1.44 µM PD-L1), Sort 3 (**c,** 1.44 nM PD-L1), Sort 4 (**d,** 144 pM PD-L1), Sort 5 (**e,** 14.4 nM PD-L1 and 864 nM PD-1 competitor).

**Extended Data Figure 3.**
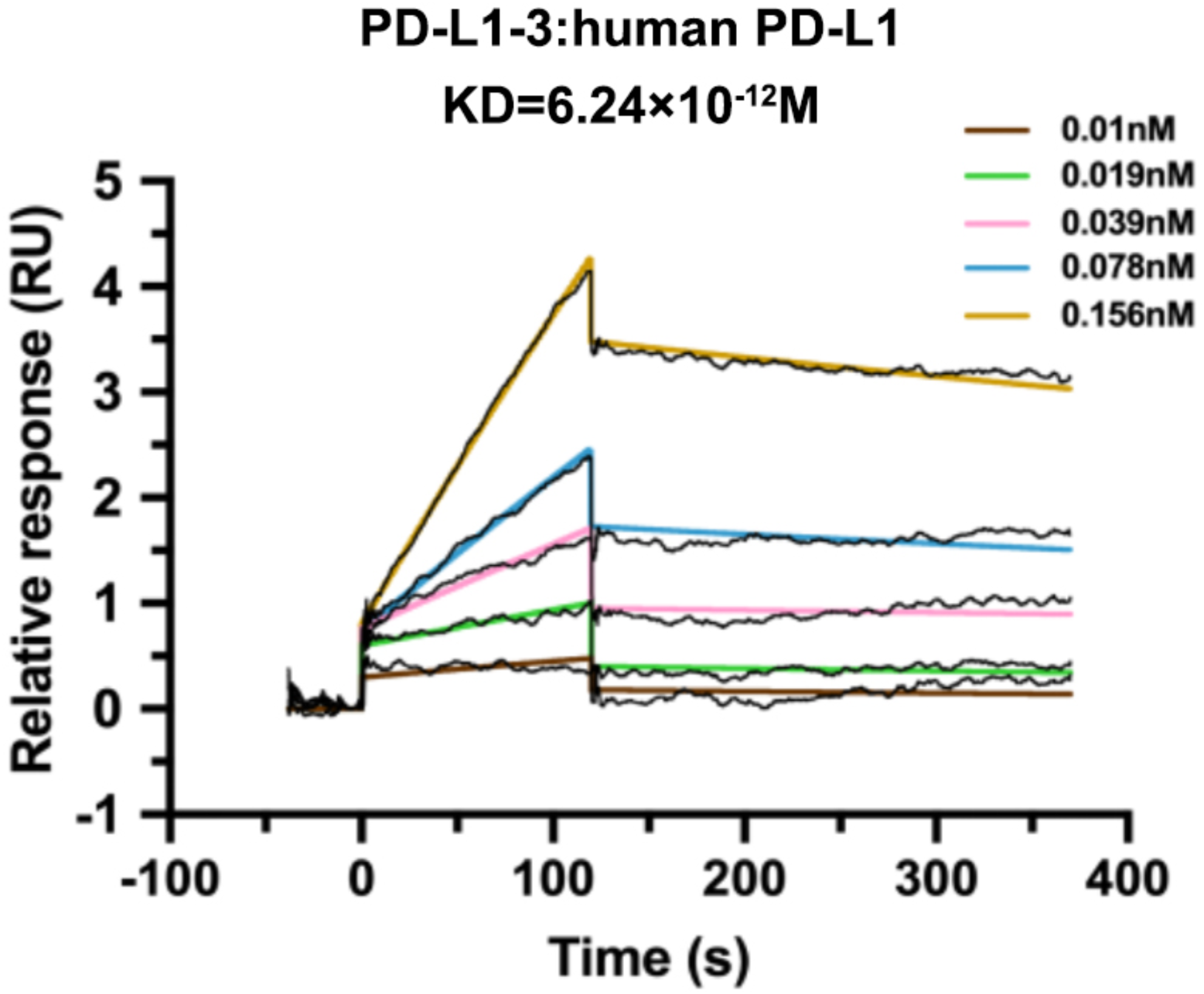
SPR analysis of PD-L1-3 binding to PD-L1. The binding affinity of PD-L1-3 for human PD-L1 was measured using SPR. Following immobilization of human PD-L1 on a CM5 sensor chip, serial dilutions of PD-L1-3 (0.156-0.01 nM) in HBS-EP running buffer were analyzed. Colored solid lines represent global fits to a 1:1 binding model, with the dissociation constant (KD) values derived from these fits. RU, relative response units.

**Extended Data Figure 4.**
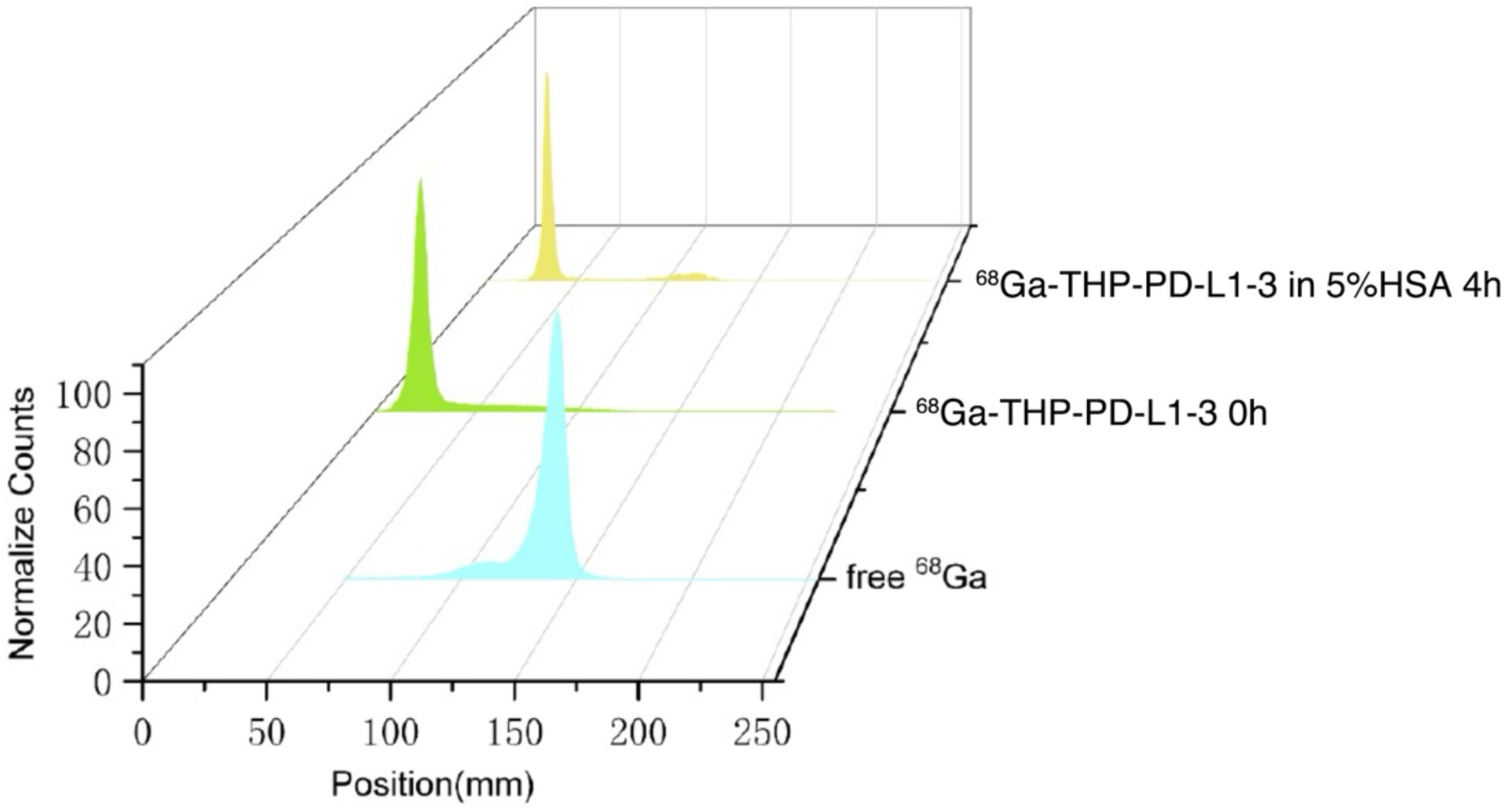
Stability assessment of [^68^Ga]Ga-THP-PD-L1-3. [^68^Ga]Ga-THP-PD-L1-3 stability in 5% human serum albumin (HSA) at 37 °C for 4 h, analyzed by radio-TLC (radiochemical purity: 96.35%), with co-chromatographed controls: free ^68^Ga and baseline samples (t=0).

**Extended Data Figure 5.**
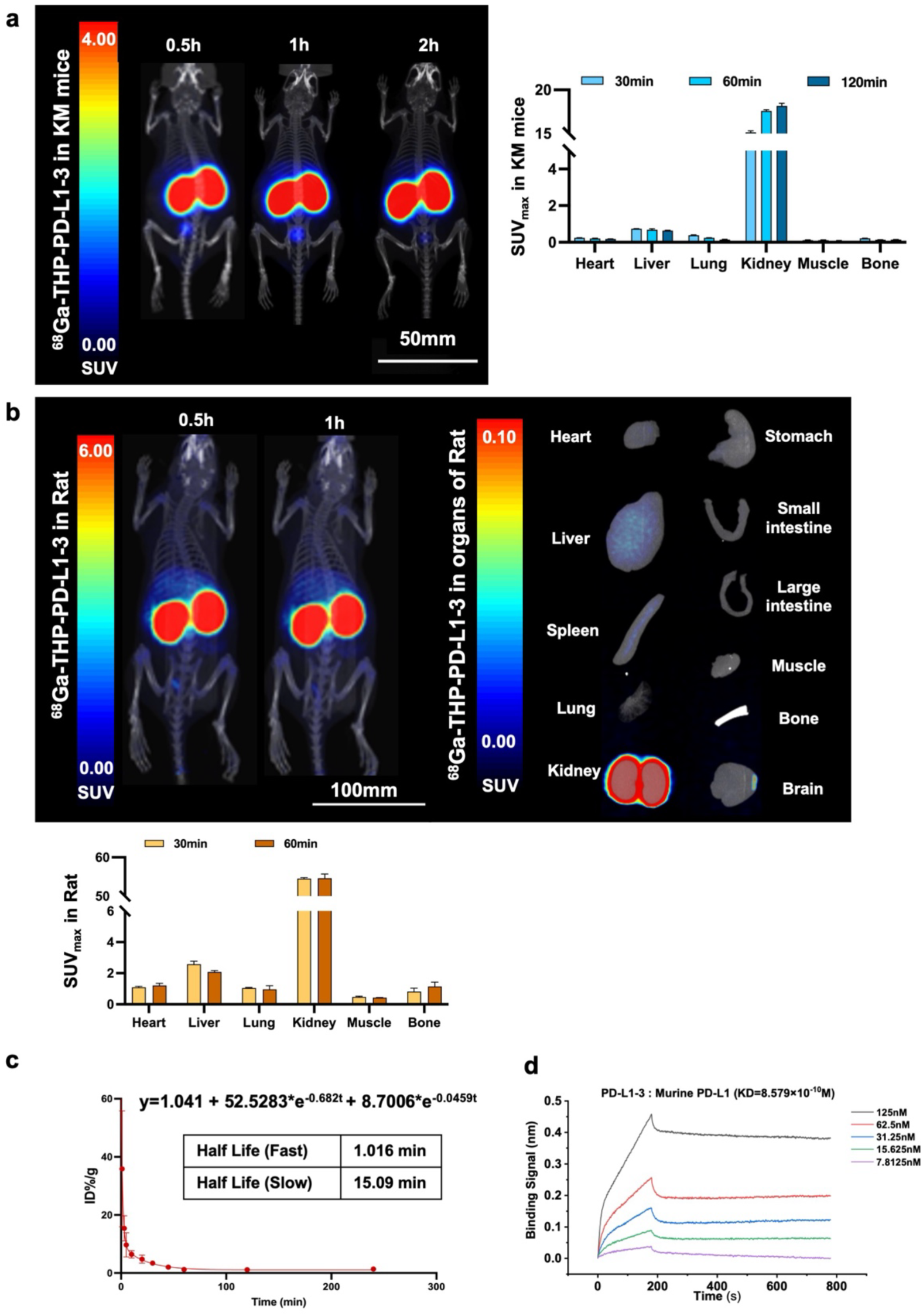
Vivo biodistribution and pharmacokinetics. **a**, PET imaging of [^68^Ga]Ga-THP-PD-L1-3 at 0.5h, 1h, and 2h post-injection in female KM mice, with SUVmax analysis of major uptake organs (n=2). **b**, PET imaging of [^68^Ga]Ga-THP-PD-L1-3 at 0.5h and 1h post-injection in female rats (n=2), with stereoscopic organ autoradiography at 2h and SUVmax analysis of major uptake organs. The biodistribution pattern in rats remained similar to that in KM mice. **c**, Blood activity-time curve of [^68^Ga]Ga-THP-PD-L1-3 in normal KM mice.

**Extended Data Figure 6.**
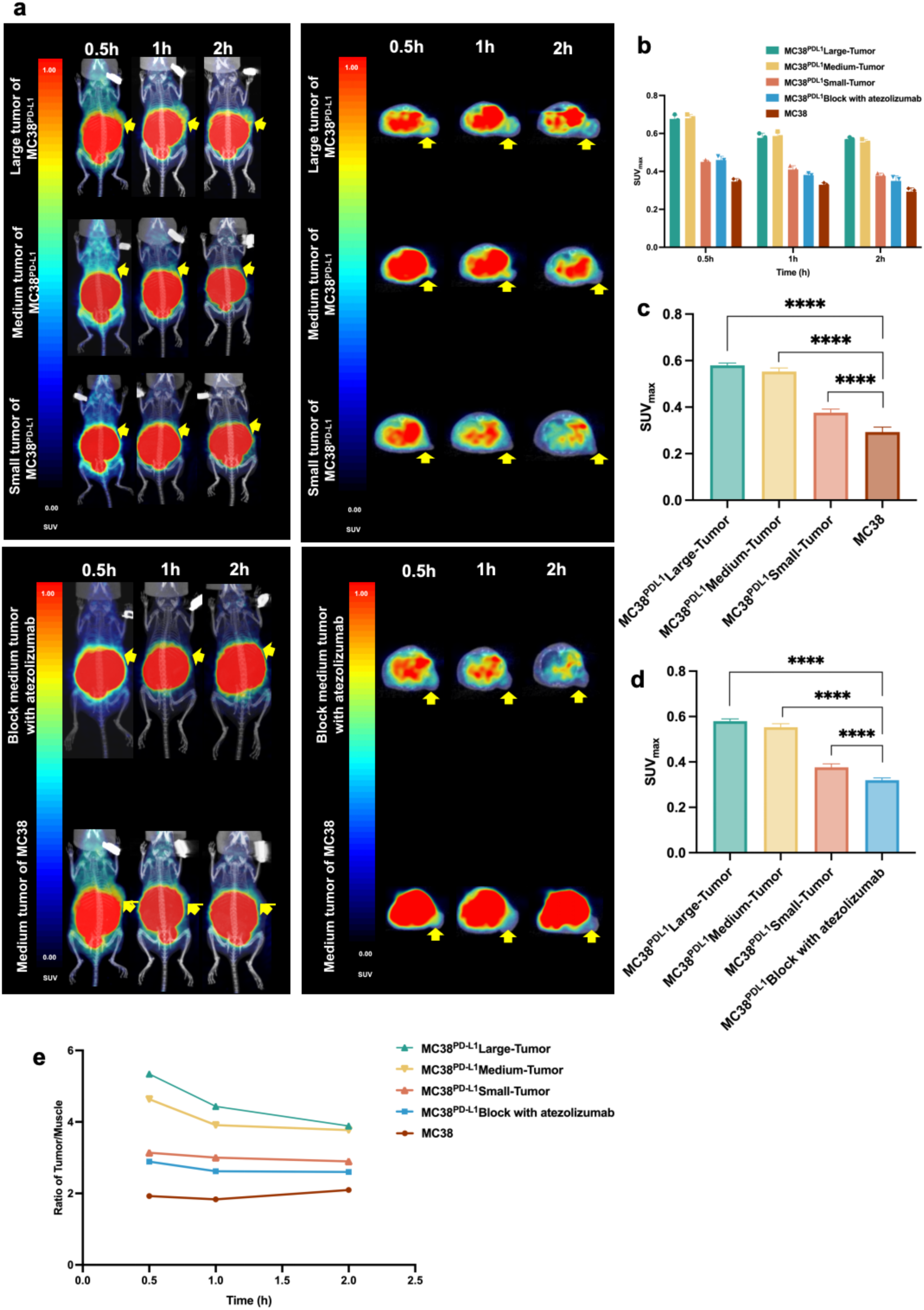
PET/CT imaging showing biodistribution of [^68^Ga]Ga-THP-PD-L1-3 across tumor volume gradients. **a**, Volume-stratified tumor models in C57BL/6N mice (n=3/group) with PET/CT at 0.5, 1, 2 h + atezolizumab blocking (2 mg). **b**, Cohort-specific SUVmax at 0.5 h, 1 h, and 2 h. **c**, Tumor uptake ([^68^Ga]Ga-THP-PD-L1-3) in volume-stratified MC38^PD-L1^ vs MC38 tumors at 2 h. **d**, Blocking effect on uptake ([^68^Ga]Ga-THP-PD-L1-3) at 2 h (mean ± SD; ****P<0.0001, **P=0.0058). **e**, Tumor-to-muscle ratio comparison at 0.5 h, 1 h, and 2 h.

**Extended Data Figure 7.**
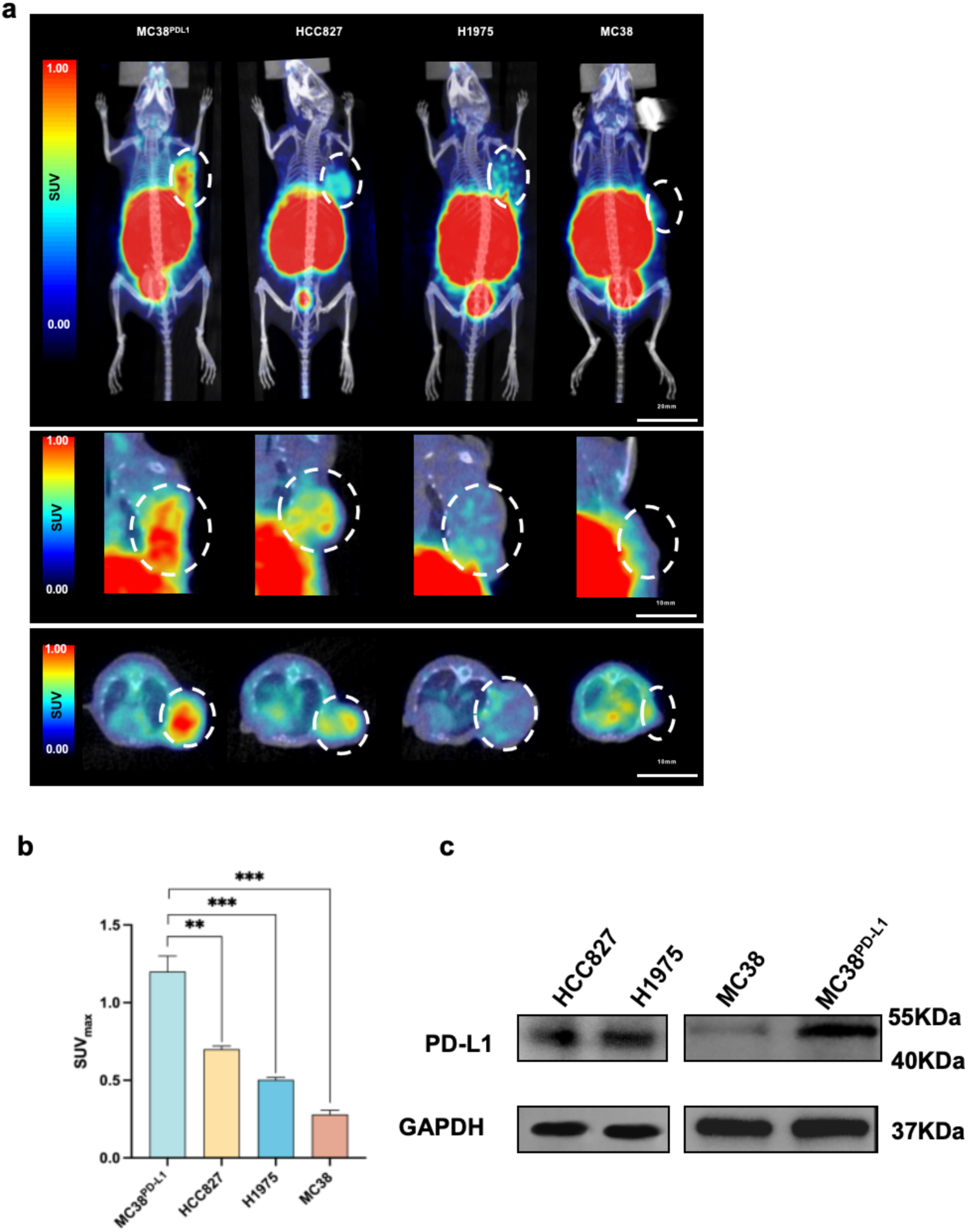
PET/CT imaging demonstrating [^68^Ga]Ga-THP-PD-L1-3 biodistribution across PD-L1 expression gradients. **a**, micro-PET at 0.5 h in tumor models (MC38^PD-L1^, HCC827, H1975, MC38). **b**, Tumor SUVmax by model (***(P<0.0005), **(P<0.005) vs. MC38; unpaired t-test). **c**, PD-L1 expression validation by Western blot (MC38^PD-L1^, HCC827, H1975, MC38).

**Extended Data Figure 8.**
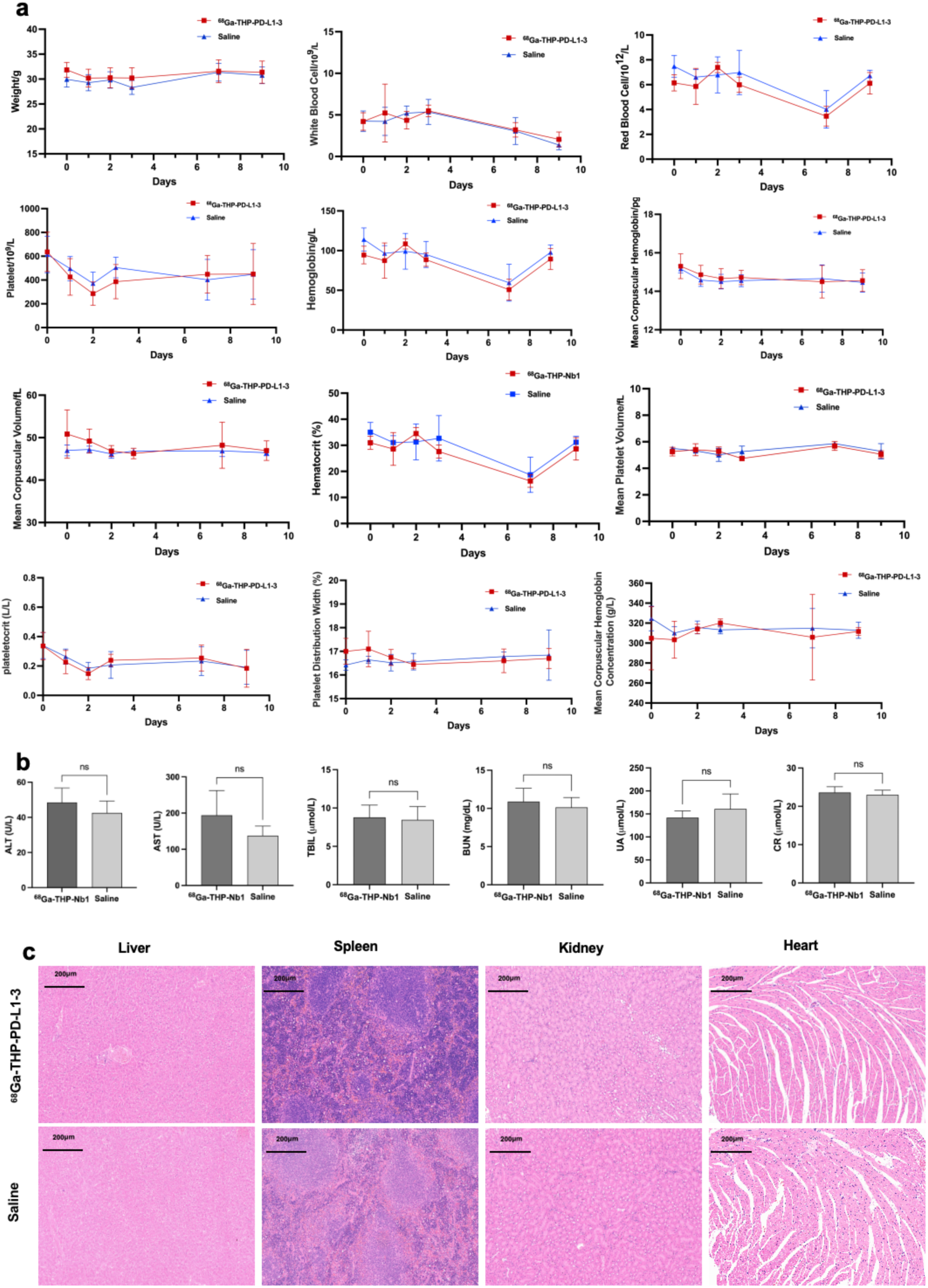
The acute toxicity assessment of [^68^Ga]Ga-THP-PD-L1-3. KM mice were randomly allocated into control and experimental groups (n=6 per group). The experimental group received a single intravenous dose of [^68^Ga]Ga-THP-PD-L1-3 (18.5 MBq in 200 μL), while the control group was administered an equivalent volume of saline. **a,** Routine blood test results of BALB/c female mice in the radiotoxicity study of [^68^Ga]Ga-THP-PD-L1-3 (n=6). **b,** Blood biochemical examination results of BALB/c female mice in the radiotoxicity study of [^68^Ga]Ga-THP-PD-L1-3 (n=6). Error bars indicate the mean ± SD. Comparison between groups was calculated using the unpaired t-test. ns, not significant. **c,** H&E staining results of major organs taken from the experimental group and control group.

**Extended Data Figure 9.**
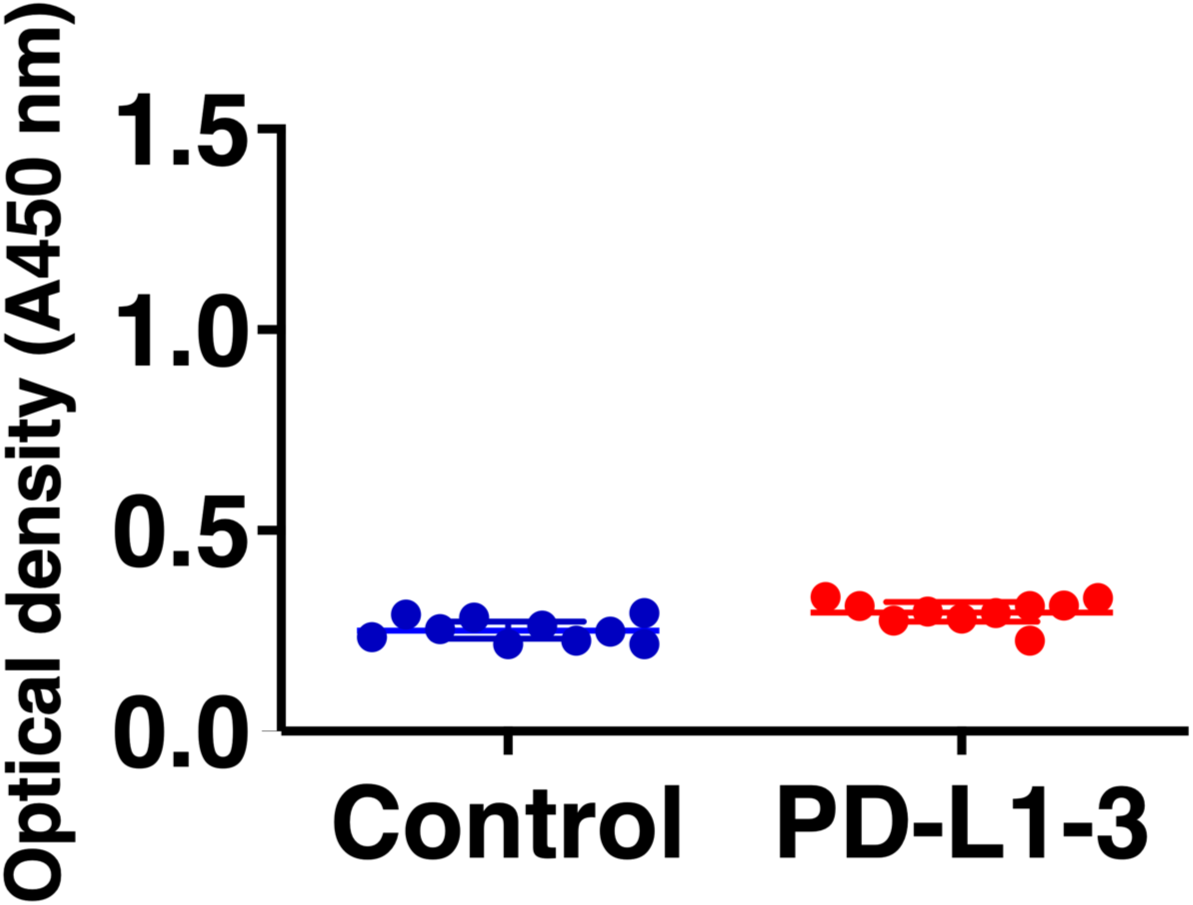
Immunogenicity of PD-L1-3. C57BL/6 mice (n=6 per group) were treated every 72 hours with 50 µg of PD-L1-3 or control binder by intraperitoneal (i.p.) administration. On day 18 post-treatment, blood samples were collected and anti-PD-L1-3 antibodies were measured by ELISA (mean ± SEM, three independent experiments).

**Extended Data Figure 10.**
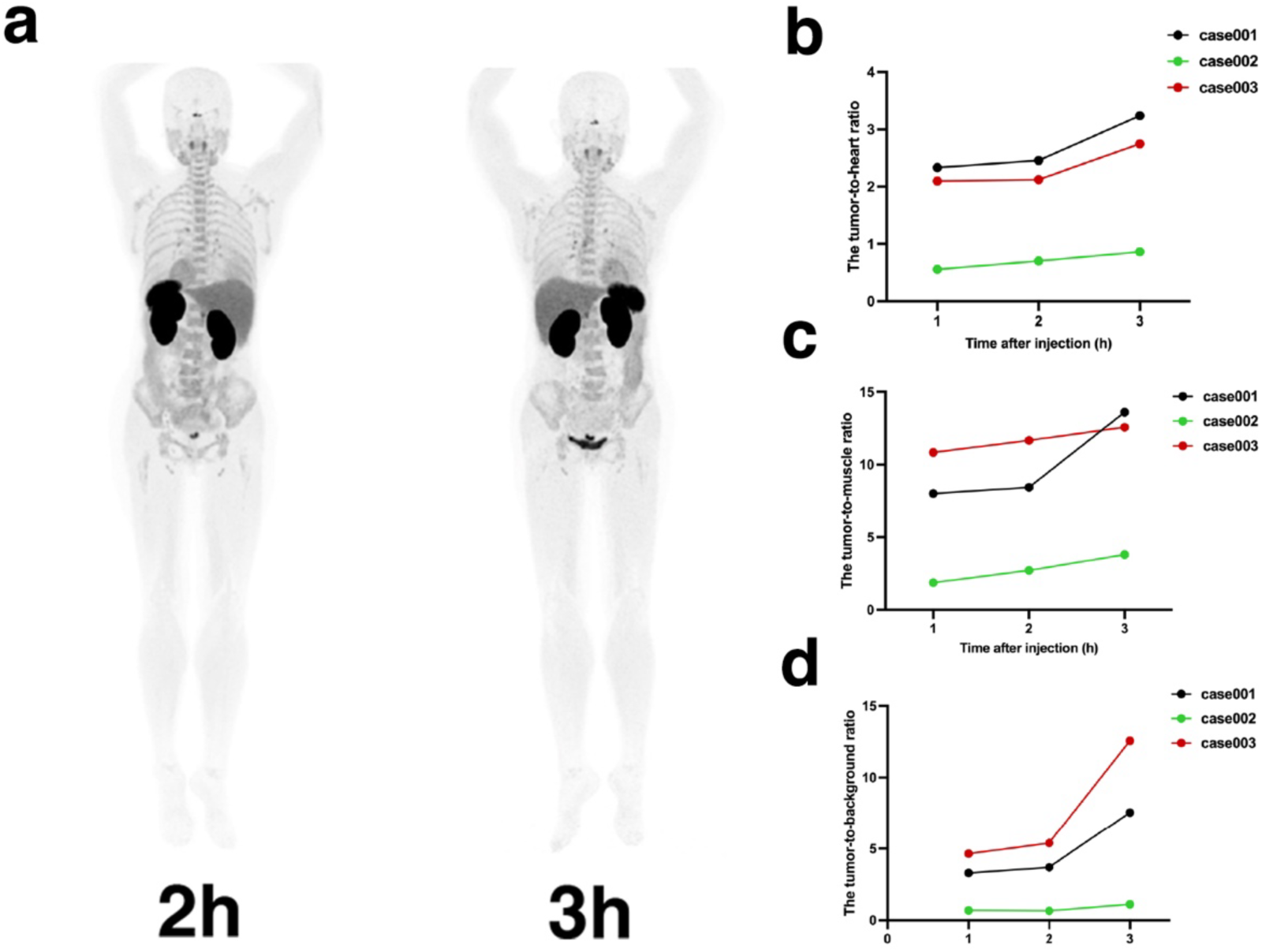
PET imaging of [^68^Ga]Ga-THP-PD-L1-3 in a patient with CSCC. **a**, Maximum intensity projection (MIP) imaging was conducted in the same patient at 120 min and 180 min post injection.

**Extended Data Figure 11.**
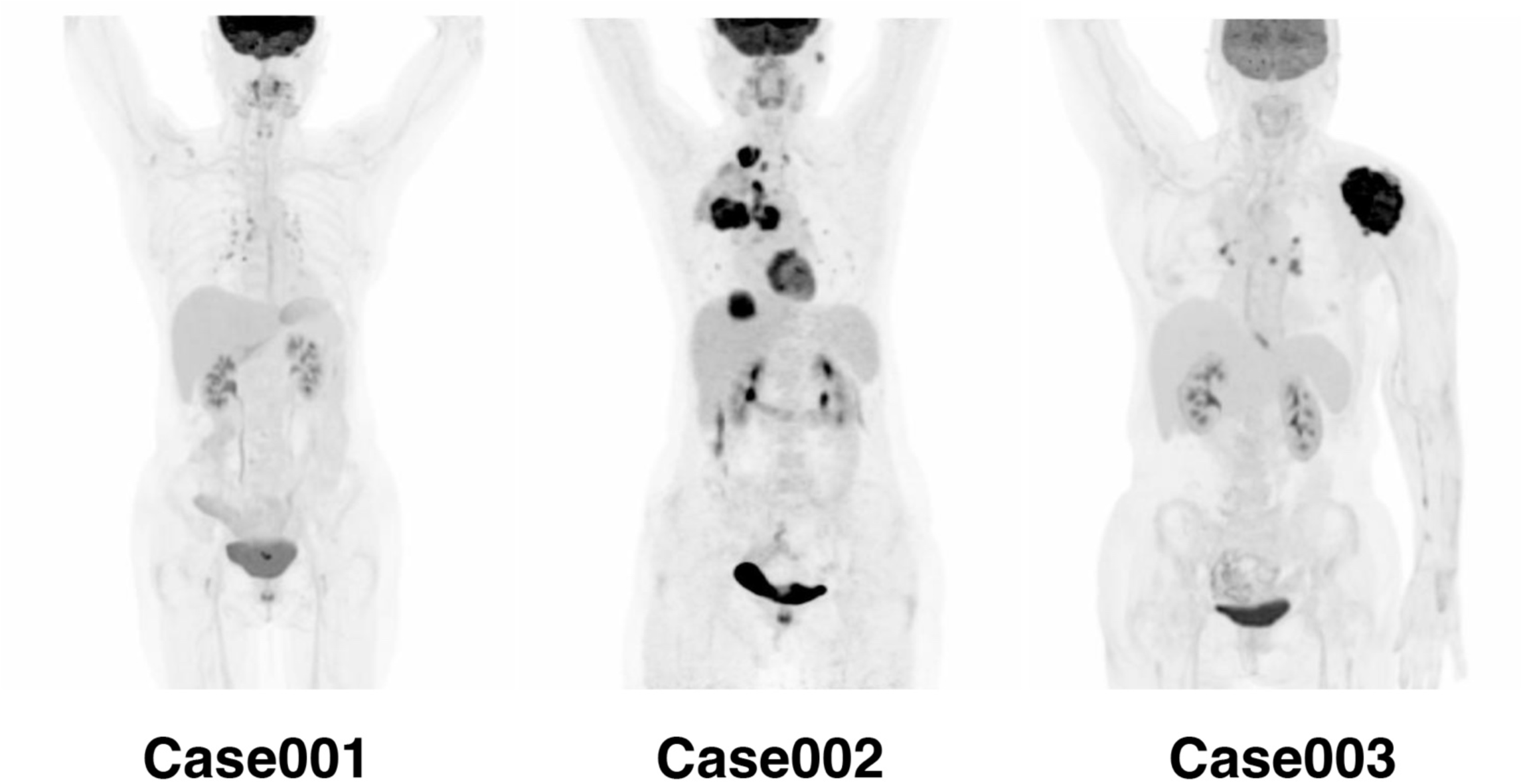
^18^F-FDG PET/CT imaging with maximum intensity projection (MIP) reconstruction of three patients.

**Extended Data table 1.**
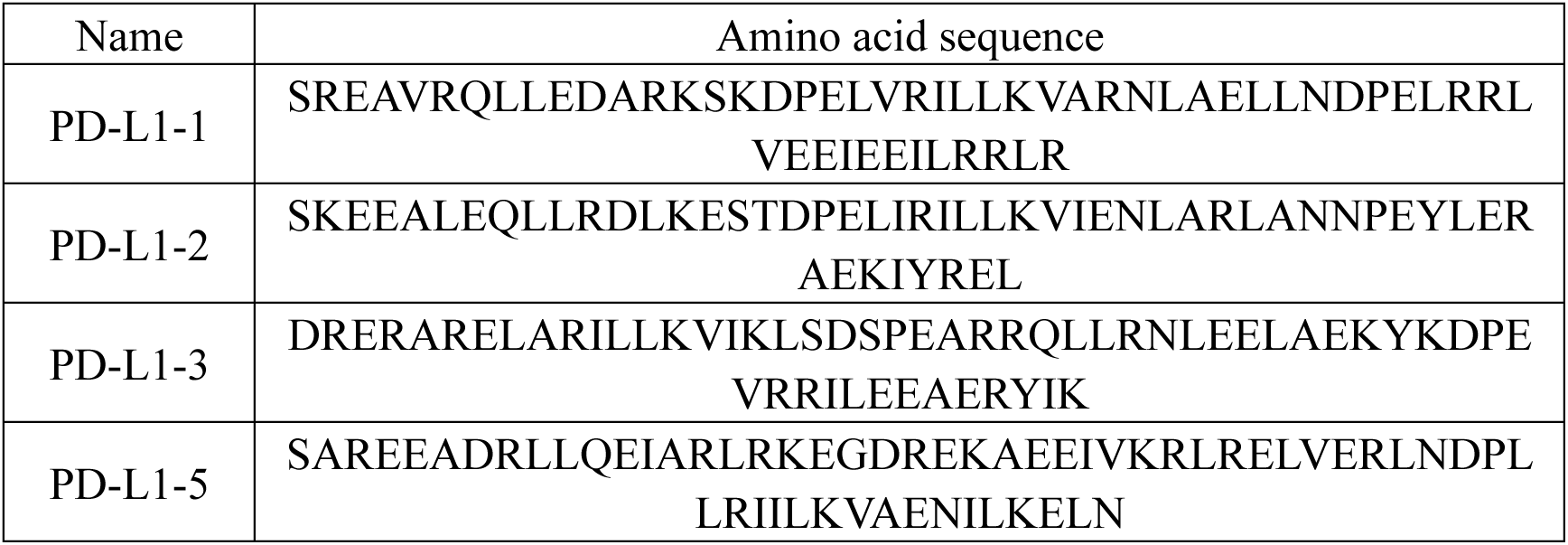
The sequences of all biochemically characterized binder proteins.

**Extended Data table 2.** Crystallographic data collection and refinement statistics.

|  | **PD-L1-3/PD-L1** |
| --- | --- |
| **Resolution range** | 30.46 - 1.77 (1.833 - 1.77) |
| **Space group** | P 1 |
| **Unit cell** | 44.872 58.956 63.299 92.515 90.015 94.804 |
| **Total reflections** | 165257 (16908) |
| **Unique reflections** | 60509 (6050) |
| **Multiplicity** | 2.7 (2.8) |
| **Completeness (%)** | 96.04 (95.56) |
| **Mean I/sigma(I)** | 4.26 (2.12) |
| **Wilson B-factor** | 16.20 |
| **R-merge** | 0.1528 (0.543) |
| **R-meas** | 0.1891 (0.6735) |
| **R-pim** | 0.1096 (0.3927) |
| **CC1/2** | 0.97 (0.673) |
| **CC\*** | 0.992 (0.897) |
| **Reflections used in refinement** | 60511 (6048) |
| **Reflections used for R-free** | 2960 (284) |
| **R-work** | 0.2009 (0.2631) |
| **R-free** | 0.2746 (0.3665) |
| **CC(work)** | 0.929 (0.852) |
| **CC(free)** | 0.846 (0.595) |
| **Number of non-hydrogen atoms** | 6059 |
| **macromolecules** | 5660 |
| **ligands** | 4 |
| **solvent** | 395 |
| **Protein residues** | 688 |
| **RMS(bonds)** | 0.006 |
| **RMS(angles)** | 0.84 |
| **Ramachandran favored (%)** | 96.58 |
| **Ramachandran allowed (%)** | 3.42 |
| **Ramachandran outliers (%)** | 0.00 |
| **Rotamer outliers (%)** | 1.96 |
| **Clashscore** | 6.45 |
| **Average B-factor** | 21.57 |
| **macromolecules** | 20.77 |
| **ligands** | 52.68 |
| **solvent** | 32.73 |

Statistics for the highest-resolution shell are shown in parentheses.

**Extended Data table 3.** Patient Characteristics.

| **Patient NO.** | **Gender** | **tumor type\*** | **ECOG score** | **PD-L1 Expression** | **[<sup>68</sup>Ga]Ga-THP-PD-L1-3 SUV<sub>max</sub> 2h/3h** | **FDG PET SUV<sub>max</sub>** |
| --- | --- | --- | --- | --- | --- | --- |
| Case 1 | Female | CSCC | 0 | 70 *** | 7.0/8.8 | 10.2 |
| Case 2 | Female | LUAC | 0 | 1 %** | 1.4/1.4 | 17.4 |
| Case 3 | Female | LUAC | 1 | 95 %** | 5.9/6.8 | 15.0 |

\* LUAC, Lung adenocarcinoma; CSCC, Cervical squamous cell carcinoma
\*\* TPS, tumor proportion score
\*\*\* CPS, Combined positive score

**Extended Data table 4.** [^68^Ga]Ga-THP-PD-L1-3 treatment-related adverse events in 3 evaluable patients.

|  | No. (%) of events<br>Any grade | Grade 3* |
| --- | --- | --- |
| Pruritus | - | - |
| Alanine aminotransferase<br>increased | - | - |
| Alkaline phosphatase<br>increased | - | - |
| Alopecia | - | - |
| Aspartate aminotransferase<br>increased | - | - |
| Anorexia | - | - |
| Arthralgia | - | - |
| Chills | - | - |
| Diarrhea | - | - |
| Dizziness | - | - |
| Dry eyes | - | - |
| Dry mouth | - | - |
| Dry skin | - | - |
| Edema | - | - |
| QT corrected interval<br>prolonged | - | - |
| Fatigue | - | - |
| Flatulence | - | - |
| Flu like symptoms | - | - |
| Hyperthyroidism | - | - |
| Hypothyroidism | - | - |
| Gammaglutamyltransferase<br>increased | - | - |
| Infusion related reaction | - | - |
| Insomnia | - | - |
| Myalgia | - | - |
| Nausea | - | - |
| Pain in extremity | - | - |
| Paresthesia | - | - |
| Platelet count decreased | - | - |
| Pneumonitis | - | - |
| Pruritus | - | - |
| Rash | - | - |

\* No grade 4 events were observed. No, number.

